# An explainable machine learning-based phenomapping strategy for adaptive predictive enrichment in randomized controlled trials

**DOI:** 10.1101/2023.06.18.23291542

**Authors:** Evangelos K Oikonomou, Phyllis M. Thangaraj, Deepak L Bhatt, Joseph S Ross, Lawrence H Young, Harlan M Krumholz, Marc A Suchard, Rohan Khera

**Author notes:** Address for correspondence: Rohan Khera, MD, MS, 195 Church St, 6^th^ Floor, New Haven, CT 06510; @rohan_khera; ORCID: 0000-0001-9467-6199.

## Abstract

Randomized controlled trials (RCT) represent the cornerstone of evidence-based medicine but are resource-intensive. We propose and evaluate a machine learning (ML) strategy of adaptive predictive enrichment through computational trial phenomaps to optimize RCT enrollment. In simulated group sequential analyses of two large cardiovascular outcomes RCTs of (1) a therapeutic drug (pioglitazone versus placebo; Insulin Resistance Intervention after Stroke (IRIS) trial), and (2) a disease management strategy (intensive versus standard systolic blood pressure reduction in the Systolic Blood Pressure Intervention Trial (SPRINT)), we constructed dynamic phenotypic representations to infer response profiles during interim analyses and examined their association with study outcomes. Across three interim timepoints, our strategy learned dynamic phenotypic signatures predictive of individualized cardiovascular benefit. By conditioning a prospective candidate’s probability of enrollment on their predicted benefit, we estimate that our approach would have enabled a reduction in the final trial size across ten simulations (IRIS: – 14.8% ± 3.1%, *p_one-sample t-test_*=0.001; SPRINT: –17.6% ± 3.6%, *p_one-sample t-test_*<0.001), while preserving the original average treatment effect (IRIS: hazard ratio of 0.73 ± 0.01 for pioglitazone vs placebo, vs 0.76 in the original trial; SPRINT: hazard ratio of 0.72 ± 0.01 for intensive vs standard systolic blood pressure, vs 0.75 in the original trial; all with *p_one-sample t-test_*<0.01). This adaptive framework has the potential to maximize RCT enrollment efficiency.

## INTRODUCTION

Large randomized controlled trials (RCTs) represent the cornerstone of evidence-based medicine and are the scientific and regulatory gold standard.^1,2^ Despite their strengths, they are often both resource and time intensive.^3^ The required investments are particularly large for RCTs evaluating the effects of novel therapies on major clinical endpoints such as mortality or acute cardiovascular events among patients with chronic cardiometabolic or other disorders.^4–6^ Modern pivotal trials have been facing exponentially rising costs as more patients and clinic visits are needed to prove a treatment effect, with the median cost per approved drug estimated at $48 million and a median cost of $41413 per patient enrolled. Across studies, the largest single factor driving cost was the number of patients required to establish the treatment effects.^5,6^ With a growing pipeline of potential new therapeutics, there is a need to explore alternative methods of conducting RCTs to increase their efficiency and provide high-quality evidence to ensure patient safety and regulatory compliance.^7,8^

We have recently described a machine learning (ML) method that leverages the phenotypic diversity of patients in RCTs and the random allocation of the intervention to define signatures of individualized treatment effects.^9–11^ Our method is based on an approach that creates a multidimensional representation of an RCT population across all pre-randomization features (“phenomap”) and extracts signatures that define consistent benefit or risk from each study arm. This approach has been validated retrospectively across several RCTs,^9–11^ however its utility in the context of an adaptive trial design has not been explored.

Adaptive trials, which allow prospectively planned modifications to the study design of clinical trials based on accumulating data from already enrolled patients,^12^ have been proposed as a potential solution.^13,14^ In a guidance statement, the United States Food and Drug Administration (FDA) proposes that “a trial might enroll subjects from the overall trial population up through an interim analysis, at which time a decision will be made based on prespecified criteria whether to continue enrollment in the overall population or to restrict future enrollment to the targeted subpopulation”.^15^ The ability to adjust the trial to new information maximizes its efficiency while ensuring safety by detecting early harm or lack of effectiveness.^12,16,17^ However, defining the ways in which to adapt a trial *a priori* remains challenging.

In the present study, we propose and evaluate an adaptive approach that uses phenomapping-derived study arm effect differences for similar patients grouped on their complex phenotypic features to design predictively enriched clinical trials. We demonstrate the application of this hypothesis using individual participant data from two large cardiovascular outcomes trials, assessing the effects of our proposed approach on efficacy, safety endpoints, as well as the final trial composition.

## RESULTS

### Study population

The study was designed as a post hoc simulation of real-world clinical trial data from a double-blind, placebo-controlled, randomized trial of a drug (pioglitazone, as studied in the Insulin Resistance Intervention after Stroke [IRIS] trial),^18^ and a disease management strategy (intensive versus standard blood pressure reduction in the Systolic Blood Pressure Intervention Trial [SPRINT])^19^ (**Fig. 1a**). The detailed protocol, study population demographics, and results have been previously reported and are further summarized in the **Methods**.^18,19^ Briefly, IRIS included 3876 patients, 40 years of age or older, with a recent ischemic stroke or a transient ischemic attack (TIA) during the 6 months before randomization, who did not have diabetes mellitus but had evidence of insulin resistance based on a homeostasis model assessment of insulin resistance (HOMA-IR) index score of 3.0 or greater. Participants were randomly assigned in a 1:1 ratio to receive either pioglitazone or a matching placebo. Enrollment occurred over 2899 days (7.9 years), from February 2005 until January 2013, with the final study report including 3876 patients (median age 62 [IQR 55-71] years, n=1338 (34.5%) women). Participants were followed for a median of 4.7 [IQR 3.2-5.0] years for the primary endpoint of fatal or non-fatal stroke or myocardial infarction, which occurred in 175 of 1939 (9.0%) participants in the pioglitazone group and 228 of 1937 (11.8%) participants in the placebo group.

**Fig. 1.**
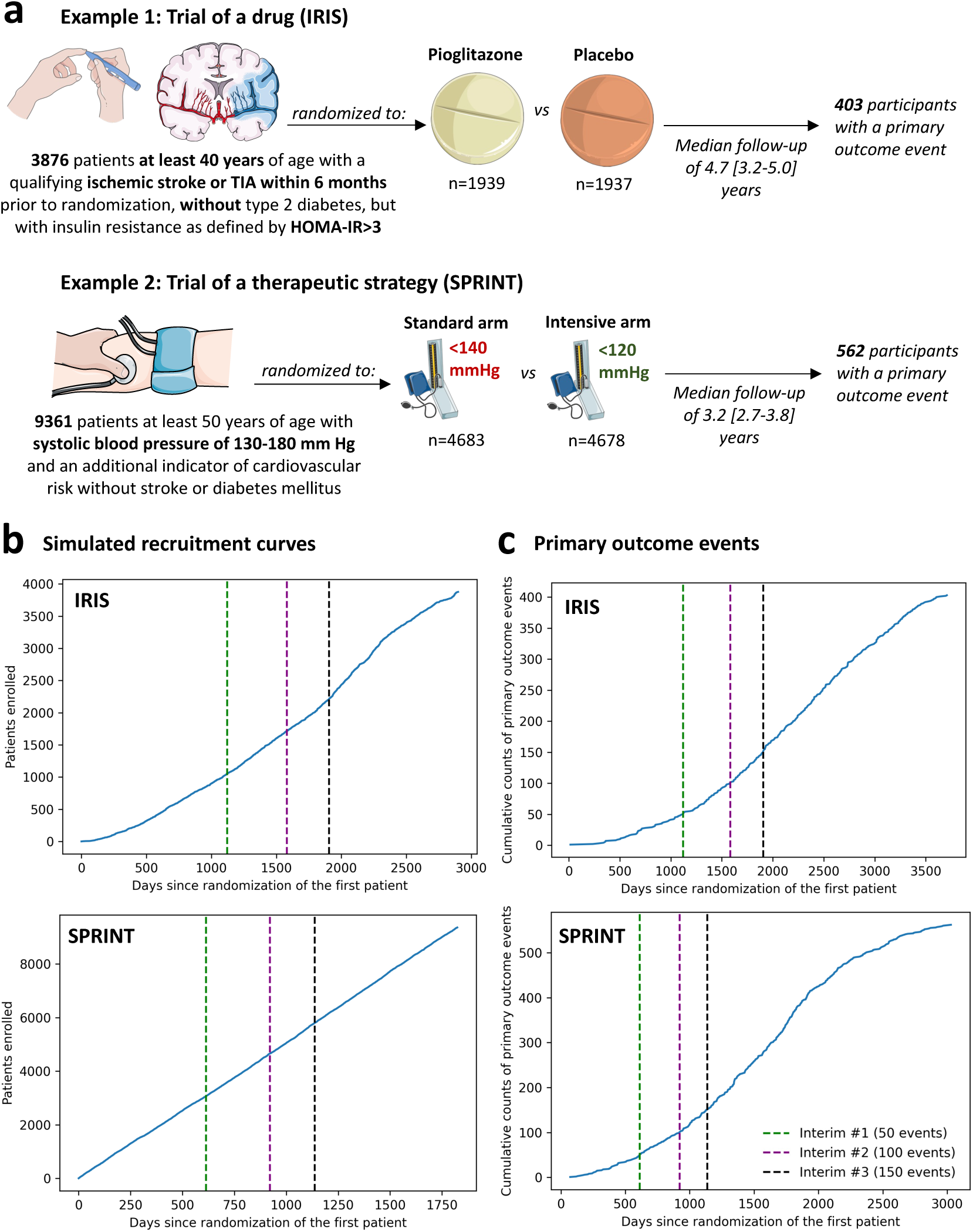
| Summary of the study design. (**a**) Visual summary of the original IRIS and SPRINT trials. **(b)** Cumulative recruitment & **(c)** primary outcome event numbers across different simulated trial timepoints. The vertical dashed lines denote the interim analysis timepoints (green: first interim timepoint; purple: second interim analysis timepoint; black: third interim analysis timepoint). HOMA-IR: homeostasis model assessment of insulin resistance; IRIS: Insulin Resistance Intervention after Stroke; SPRINT: Systolic Blood Pressure Intervention Trial; TIA: transient ischemic attack.

The SPRINT trial enrolled 9361 participants (median age 67 [61-76] years, n=3332 [35.6%] women) with a systolic blood pressure (SBP) of 130-180 mm Hg as well as an additional indicator of cardiovascular risk, with random assignment to targeting an SBP of less than 120 mm Hg (intensive treatment arm) versus less than 140 mm Hg (standard treatment arm). Patients with diabetes mellitus, prior stroke, or dementia were excluded. To simulate a longer enrollment period similar to IRIS, we modeled a steady recruitment rate over 5 years (vs 2.4 years in the original study) and used the original follow-up data with a median period of 3.2 [IQR 2.7-3.8] years for the primary composite outcome of time-to-first myocardial infarction/acute coronary syndrome, stroke, acute decompensated heart failure, or cardiovascular mortality. This occurred in 243 of 4678 (6.8%) participants in the intensive and 319 of 4683 (5.2%) participants in the standard arm.

### Defining a group sequential trial design

We defined a group sequential design, with three total interim analyses planned before the final analysis, which was performed once all primary events had occurred in the original trial. Each analysis examined intervention superiority, assuming a power of 80% and a significance level of 0.025 for one-sided test,^20^ providing adjusted significance levels (alpha) at each analysis timepoint based on the O’Brien-Fleming and alpha-spending Pocock methods (**Supplementary Table 1**). For IRIS, we assumed that the primary outcome would occur in 11.8% vs 9% of the placebo– and pioglitazone-treated arms. This was based on the observed outcomes given that amendments were to the original trial protocol during its course. For SPRINT, we assumed the respective primary outcome would occur in 6.8% vs. 5.4% of the standard and intensive arms, per the original power calculations.^19^ We defined the interim analysis timepoints based on the occurrence of the first 50, 100 and 150 primary outcome events in the original trial. In IRIS, this corresponded to 1121, 1582, and 1905 days after the first patient was randomized. In the simulated SPRINT analysis, this corresponded to 613 days, 921 days, and 1137 days after the patient was randomized (**Fig. 1b-c**) in the trial. In both trials, all interim analyses were performed during periods of active trial enrollment.

### Learning machine learning, phenomapping-derived signatures of treatment benefit

At each of the 3 interim analyses, we adapted and implemented an ML algorithm that learned signatures of individualized treatment response for the intervention (pioglitazone, intensive SBP reduction) relative to the control arm based on data available at that time. The algorithm (**Fig. 2**) is described in detail in the **Methods**, and is based on our prior work.^9–11^

**Fig. 2.**
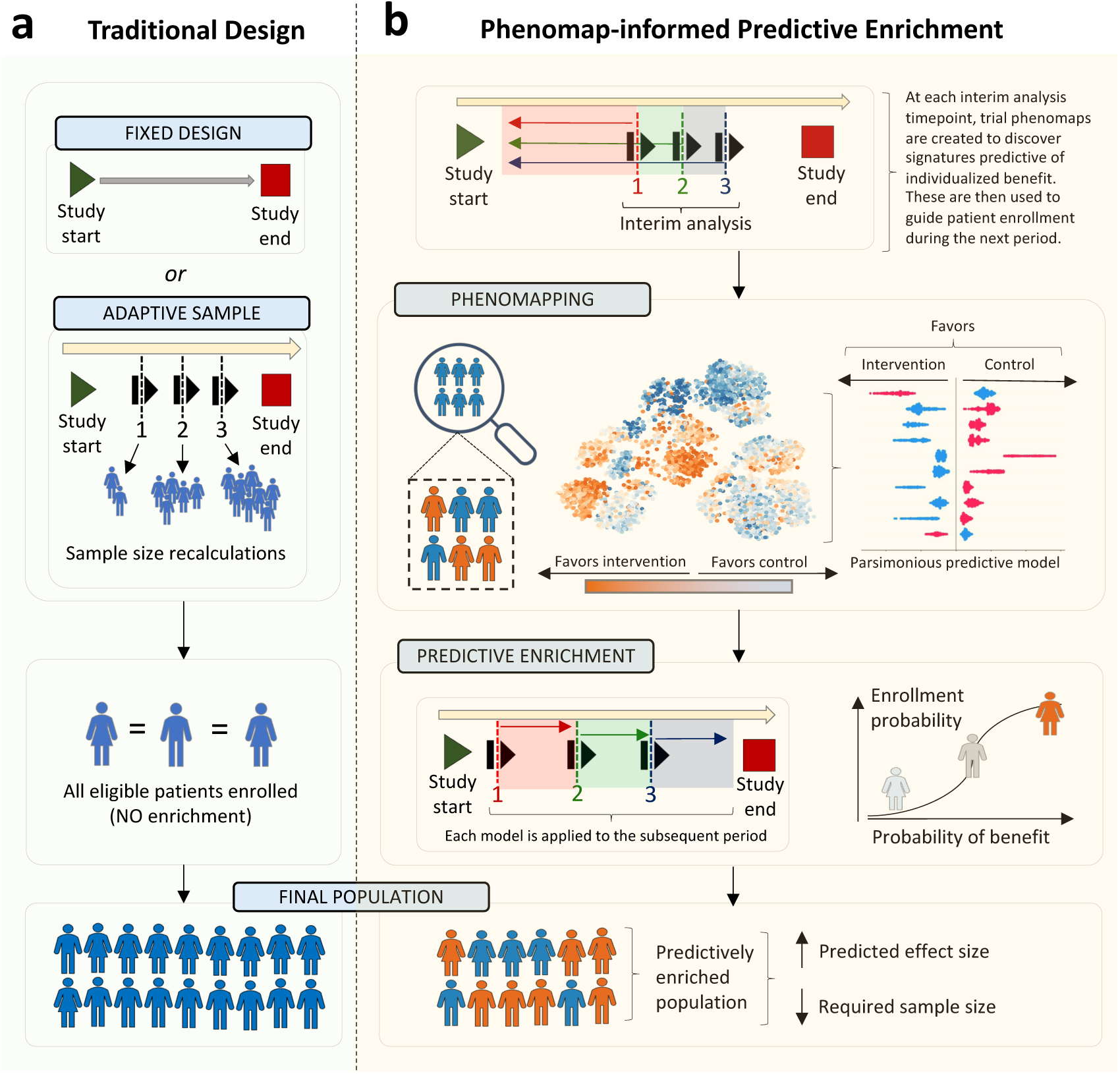
| Overview of the algorithm. (**a**) Traditional clinical trials commonly use a fixed design approach, with a pre-defined total sample size goal and inclusion criteria. In some cases, interim analyses are pre-defined to adaptively modify the target study size based on revised power calculations, however without modification of the target population. **(b)** We propose an approach of adaptive predictive enrichment through phenomapping-derived signatures of individualized benefit. At each pre-defined interim analysis timepoint, the observations and events collected up until that point are randomly split into a training and testing set. In the training set, a trial phenomap is created that represents a representation of the phenotypic similarities across all recorded baseline features. This allows the estimation of weighted average treatment effects by analyzing the observed outcomes from the phenotypic angle of each individual participant. This is followed by training of an extreme gradient boosting algorithm that links pre-randomization features to the observe treatment effect heterogeneity. If there is evidence of possible heterogeneity, power calculations are revised based on the observed effect sizes and, assuming the required final sample is lower than the originally estimated one, predictive enrichment occurs over the following period. During this time, the probability of enrollment for each prospective candidate is conditioned on their estimated benefit, whereas treatment assignment remains completely randomized. The process is repeated at each interim analysis timepoint, where a completely new model is trained.

Briefly, for each interim analysis, baseline characteristics of participants were defined based on participant assessments before randomization (summarized in **Supplementary Tables 2-3**).^11^ Participants recruited up until that stage were randomly split into training/cross-validation (50%) and testing sets (50%). In the training set, baseline data were pre-processed and used to define a phenomap, which represented the phenotypic architecture of the recruited population at that timepoint across all axes of baseline variance.

Through iterative analyses centered around each unique individual and weighted for each individual participant’s location in the phenotypic space,^11^ we defined individualized estimates of the effects of the studied intervention, as compared to control, for the primary outcome. Subsequently, using extreme gradient boosting (XGBoost) and the Boruta SHAP (Shapley additive explanations) feature selection algorithm, we built an ML framework to identify key features that collectively determined a phenotypic signature (algorithm) predictive of these individualized estimates. The predictive algorithm (relying exclusively on pre-randomization features) was then applied in the testing set to obtain ranks of predicted treatment responses based on the XGBoost regressor. To assess the stability of the predictions at each interim analysis timepoint, we performed 100 random iterations, and assessed a) how often a randomly selected patient A and patient B had consistent versus discordant relative ranks based on the XGBoost regressor (average concordance ratio); and b) the relative frequency with which individual features were selected by the Boruta SHAP method across iterations (**Supplementary** Figure 1).

We assessed for evidence of heterogeneous treatment effects (defined based on a *p* value for interaction of <0.2) by dichotomizing the population into two groups based on their predicted response, avoiding major imbalance in our subgroups by restricting the smallest group size to 20% of the population.

If there was potential evidence of heterogeneity based on this analysis, sample size calculations were updated at that interim analysis timepoint by revising the expected effect size (under the assumption of predictive enrichment) at the original power and alpha levels to ensure that predictive enrichment would maintain sufficient power at a sample size equal to or smaller than the originally planned one. We performed sample size calculations assuming various levels of predictive enrichment, which ranged from enrolling 50% to 95% of all remaining candidates, in 5% increments. If several levels met these criteria, we ultimately chose the predictive enrichment level that minimized the required sample.

Once a predictive model had been generated and our analysis in the testing set had met the criteria for possible heterogeneity in the treatment effect with sufficient power for the primary outcome, we chose to proceed with predictive enrichment. Over the subsequent period (time between the last and next interim analyses), the model was prospectively applied to all trial candidates screened after the enrichment model was identified. For example, a model trained at interim analysis timepoint #1 was applied to individuals screened between the interim analysis timepoints #1 and #2 to furnish a probability of enrollment, with all original trial participants during this period considered eligible candidates. For all candidates, the probability of being enrolled was conditioned on their predicted individualized benefit, ultimately enriching the population at the level defined during the last interim analysis sample size calculation. Alternatively, if there was no evidence of heterogeneous treatment effect, or the proposed enrichment in enrollment would not be adequately powered at a sample size equal to or lower than the originally planned one, we proceeded as originally planned and continued with standard enrollment for that period without predictive enrichment. Given the stochastic nature of the algorithm, all simulations were repeated *r*=10 times.

### Stability analysis & explainability across interim analysis timepoints

**IRIS:** In the IRIS trial, phenomapping performed at the pre-specified timepoints identified hypercholesterolemia, hypertension smoking, use of ACEi (Angiotensin-converting-enzyme inhibitors) or ARB (angiotensin receptor blockers), thiazides or beta-blockers, prior history of stroke/TIA (transient ischemic attack) and reported gender as possible predictors of treatment effect heterogeneity for pioglitazone. Across 100 stability iterations, at least one of the top 5 predictors during the first interim analysis was also present in the final interim analysis in 95% of the iterations (**Supplementary** Figures 2-3). The relative concordance ratios of predicted treatment response ranks were 1.754 [95%CI_bootstrapping_: 1.753-1.754], 1.658 [95%CI: 1.658,1.659], and 1.803 [95%CI: 1.803, 1.804] in the three sequential interim timepoints (all >1), suggesting agreement in the relative ranks across iterations.

**SPRINT:** In SPRINT, phenomapping performed at the pre-specified timepoints highlighted several features, such as a history of chronic kidney disease, aspirin use, female sex and smoking as some of the top features predicting differential responses to intensive vs standard SBP reduction. At least one of the 5 predictors during the first interim analysis was also present in the final interim analysis in 97% (97/100) iterations (**Supplementary** Figures 4-5). The relative concordance ratios of predicted treatment response ranks were 1.8391 [95% CI: 1.8371-1.8413], 2.0527 [95%CI: 2.0511-2.0543], and 1.8343 [95%CI: 1.8334,1.8352] for the 3 timepoints, all greater than 1.

### Primary (efficacy) outcomes

***IRIS:*** Across 10 simulations of the end-to-end algorithm in IRIS, we confirmed that adaptive enrichment did not impact the random assignment of the study participants to pioglitazone versus placebo, compared to the original trial (*p_chi-square test_*=1.00 across all simulations, **Supplementary Table 4).**

A strategy of selective enrollment conditioned on individualized estimates of pioglitazone benefit was associated with a significant reduction in the final sample size of (mean ± standard error of mean [SEM]) of –14.8 ± 3.1% across all simulations (3304 ± 122 vs 3876 participants in the original trial, *p_one-sample t-test_*<0.001), with point estimates of greater benefit for pioglitazone versus placebo on the primary outcome (hazard ratio [HR] estimates of 0.73 ± 0.01 vs 0.76 in the original trial) which retained statistical significance at the end of the study across all simulations (*p_one-sample t-test_* of 0.005 ± 0.001, all <0.025) (**Fig. 3**).

**Fig. 3.**
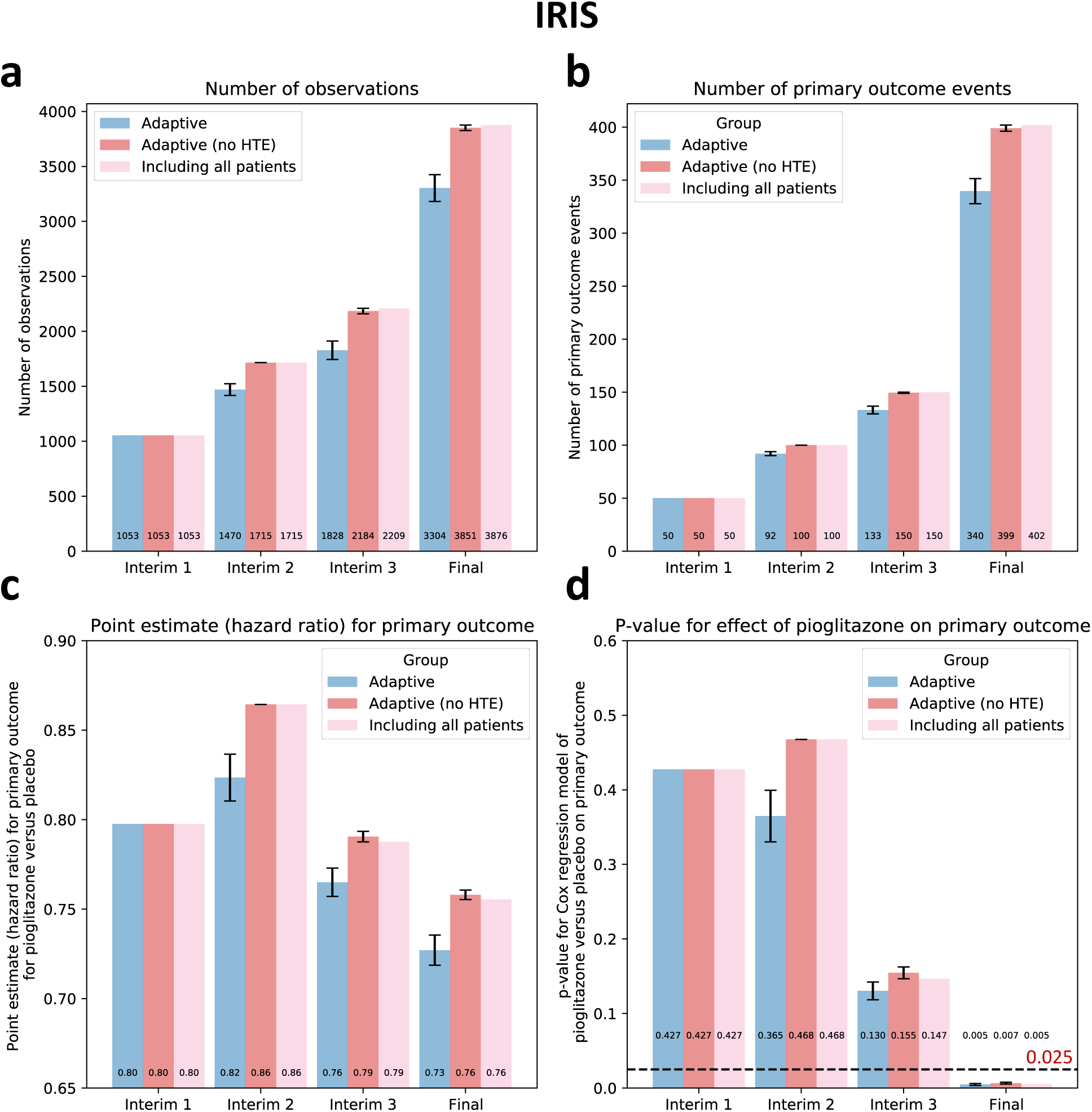
| Primary outcome results – IRIS. (**a**) Cumulative patient enrollment, **(b)** cumulative primary outcome events, **(c)** Cox regression-derived effect estimates (Hazard Ratios) for pioglitazone versus placebo, and **(d)** corresponding *p* values. For each timepoint three bars are presented; the first one on the left (blue bar) denotes the phenomapping-enriched adaptive runs, the middle (dark red) denotes an adaptive simulation after random reshuffling of baseline covariates, and the one on the right (light red) refers to the original trial. The error bars denote the standard error of mean (SEM) across n=10 adaptive simulations. Between-group comparisons and related *p* values are derived from one-sample t-test (using the original “including all patients” arm as reference). HTE: heterogeneous treatment effect; IRIS: Insulin Resistance Intervention after Stroke.

To assess the sensitivity of our approach to spurious confounding in the data, we repeated the analysis after randomly shuffling the baseline covariates of the study population. In this analysis, there was a similar average treatment effect (HR 0.76 for pioglitazone versus placebo, 95% CI 0.62-0.93) but a random association of outcomes within each study arm with the baseline covariates, thus eliminating potential heterogeneous treatment effects. As opposed to our adaptive analysis, there was no change in the final study size or primary effect estimates (*p_one-sample t-test_* =0.34 for both) (**Fig. 3**).

***SPRINT:*** Across 10 simulations in SPRINT, the random assignment of the study participants to the intensive versus standard SBP reduction arm remained balanced across all adaptive simulations and the original trial (*p_chi-square test_*=0.93, **Supplementary Table 5).**

A strategy of selective enrollment conditioned on individualized estimates of benefit from intensive versus standard SBP control was associated with a significant reduction in the final sample size of –17.6 ± 3.6% across all simulations (7711 ± 338 vs 9361 participants in the original trial, *p_one-sample t-test_*<0.001), with point estimates consistently demonstrating greater benefit from the intensive vs standard SBP reduction on the primary outcome (point HR estimates of 0.72 ± 0.01 vs 0.75 in the original trial) which retained statistical significance at the end of the study across all simulations (*p_one-sample t-test_* of 0.001±0.0003, all <0.025) (**Fig. 4**). In contrast, in a sensitivity analysis where baseline covariates were randomly shuffled within each treatment arm, we observed that our algorithm did not result in enrichment at the end of the trial, with no significant decrease in the final study size or primary effect estimates (*p_one-sample t-test_*=0.08 and 0.23, respectively).

**Fig. 4.**
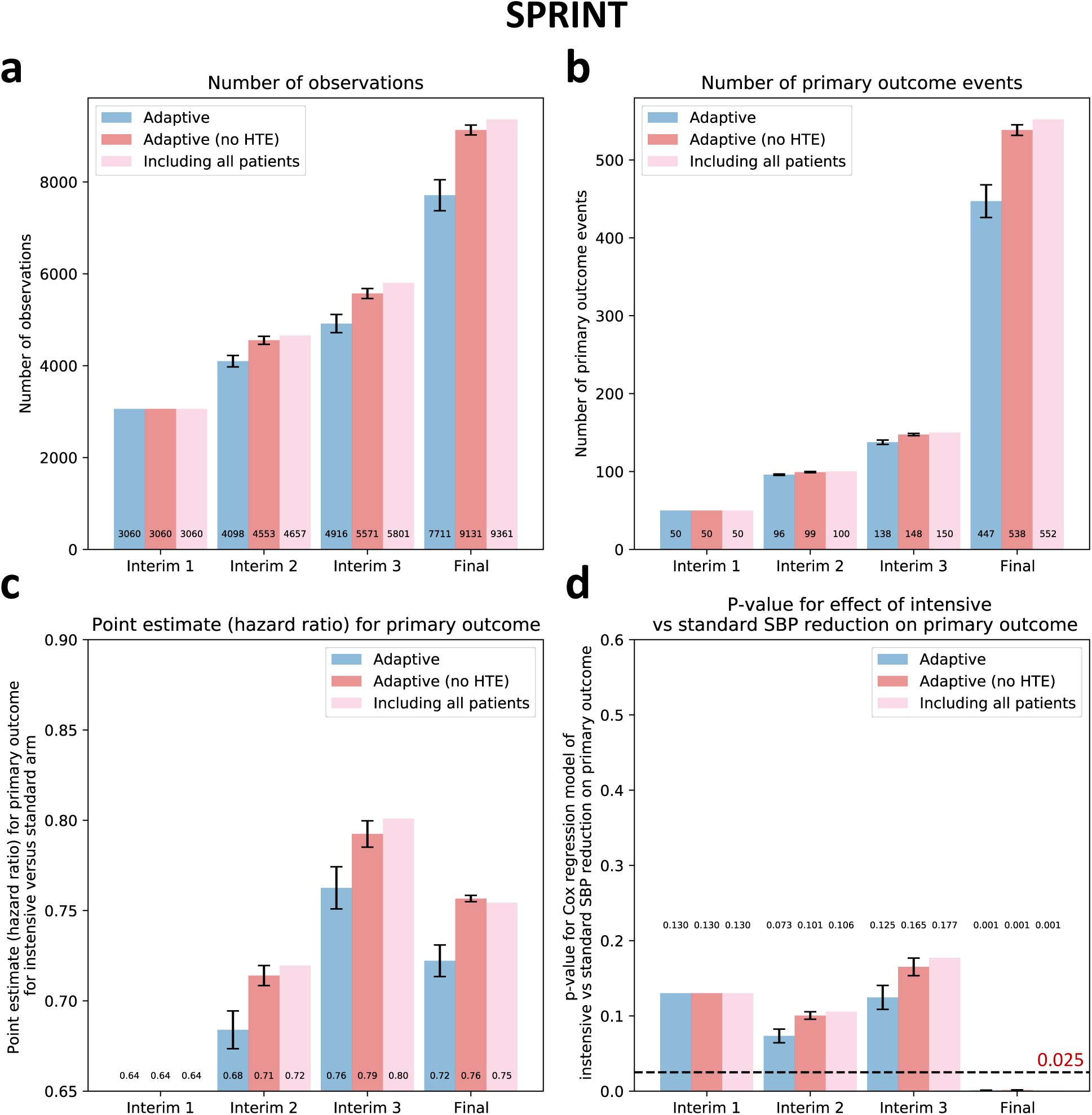
| Primary outcome results – SPRINT. (**a**) Cumulative patient enrollment, **(b)** cumulative primary outcome events, **(c)** Cox regression-derived effect estimates (Hazard Ratios) for intensive versus standard systolic blood pressure reduction, and **(d)** corresponding *p* values. For each timepoint three bars are presented; the first one on the left (blue bar) denotes the phenomapping-enriched adaptive runs, the middle (dark red) denotes an adaptive simulation after random reshuffling of baseline covariates, and the one on the right (light red) refers to the original trial. The error bars denote the standard error of mean (SEM) across n=10 adaptive simulations. Between-group comparisons and related *p* values are derived from one-sample t-test (using the original “including all patients” arm as reference). HTE: heterogeneous treatment effect; SPRINT: Systolic Blood Pressure Intervention Trial.

### Final population demographics

The distribution of the baseline demographics at the end of each simulation is summarized in **Supplementary Tables 4 and 5**. Across simulations in each trial, no key demographic population (i.e., men or women, or any specific racial/ethnic group) was excluded from the final analysis, with the representation of women ranging from 31.7% to 36.0% across simulations in IRIS (34.5% in the original trial), and 32.2% to 37.7% in SPRINT (original: 35.6%). Similarly, Black individuals formed 10.3% to 11.4% of the final population in IRIS (original: 11.0%), and 28.7% to 32.6% in SPRINT (original: 29.9%).

### Secondary (safety) outcomes

To ensure that predictive enrichment based on the projected benefit for the primary outcome is not offset by an increase in risk, we longitudinally tracked a hierarchy of key outcomes, i.e. all-cause mortality, followed by non-fatal MACE components, and then, for IRIS, hospitalization events, heart failure events, and bone fractures, and for SPRINT serious adverse events, analyzed based on the win ratio. As shown in **Fig. 5**, predictive enrichment was not associated with a significant change in the relative hazard of a hierarchical safety endpoint between the intervention and control arms, compared to the original trials.

**Fig. 5.**
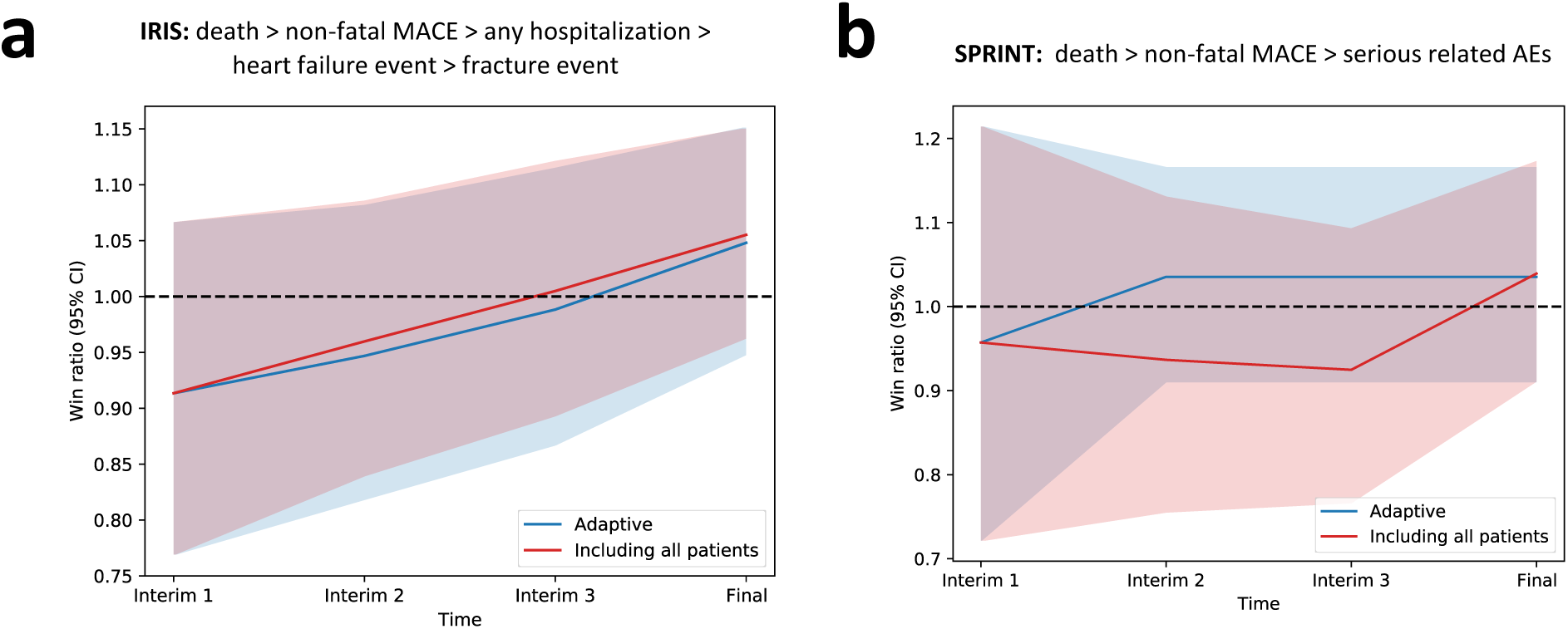
| Secondary outcome (safety) results. Win ratio with corresponding 95% confidence intervals across the pre-defined interim analysis timepoints. **(a)** Example from IRIS with outcome events ranked as follows: all-cause mortality, followed by non-fatal MACE components, and then all-cause hospitalizations, heart failure events and bone fractures; **(b)** Example from SPRINT: all-cause mortality, followed by non-fatal MACE components, and then serious adverse events. The lines correspond to the win ratio point estimate with shaded areas denoting the 95% confidence interval (for the adaptive trial design the point and upper and lower ends of the confidence interval were averaged across the ten simulations). Blue color denotes the adaptive runs, and red color denotes the original trial that includes all patients. IRIS: Insulin Resistance Intervention after Stroke; SPRINT: Systolic Blood Pressure Intervention Trial; TIA: transient ischemic attack.

### Absolute versus relative risk enrichment

To explore whether adaptive enrichment was associated with preferential enrollment of individuals at higher risk of the primary outcome, we performed subgroup analyses stratified based on the simulation strategy and the period during which a patient was enrolled. Except in one of the ten simulations after the last interim analysis timepoint in SPRINT where *p_log-rank_* was 0.04 (reference: original population), we observed no differences in the primary outcome event rates between the original and adaptive simulations (**Supplementary** Figures 6 **& 7**). This finding may suggest that predictive enrichment focuses on the effect of therapy as opposed to the underlying risk of participants.

## DISCUSSION

We present an ML-driven algorithm for adaptive predictive enrichment in RCTs that uses phenome-wide, computational trial maps to define individualized signatures of treatment benefit. Using participant-level data from the IRIS and SPRINT trials, we find that these signatures can adaptively modify the enrollment strategy in two independent trials without selectively eliminating any traditional demographic or clinical groups from the final population. We demonstrate across multiple simulations that this approach adaptively enriches for individuals most likely to benefit from the intervention and reduces the required sample size up to 18% while preserving the overall efficacy and safety signal of the intervention. These findings propose a new paradigm to maximize the efficiency and safety of clinical trials through dynamic data-driven inference.

The importance of maximizing the efficiency of clinical trial design through innovative methods has been recently embraced by numerous societies and agencies, including the FDA,^15^ and the European Medicines Agency (EMA).^21^ Adaptive trials describe a study design that permits flexibility in various aspects of the trial process, from sample size to enrollment criteria and treatment schedules, based on accumulating trial data.^16^ The tenet of this approach is that trial adaptation can minimize the size and costs of a study, as well as the potential risk to trial participants. Enrichment strategies, which are often integrated into adaptive trial designs, steer a trial toward a patient population that is most likely to respond to a given treatment.^16,22,23^ Use of these methods has been proposed across various conditions, including heart failure with preserved ejection fraction,^24^ neurodegenerative & psychiatric conditions,^25,26^ or kidney disease.^27^

To improve the precision of RCTs, prior studies have implemented various approaches. For instance, risk-based *prognostic enrichment* relies on the selective recruitment of individuals at high risk of a given condition, thus increasing the statistical power of a study for a given sample size and the chance of demonstrating a high absolute treatment effect. This can be achieved through risk algorithms,^23,25,28,29^ and imaging or circulating biomarkers with prognostic value.^22,30–32^ This approach, however, may affect the generalizability of a trial’s findings and does not evaluate whether treatment allocation to specific patient phenotypes aligns with those most likely to benefit. To address this issue, *predictive enrichment* focuses on the individualized effects of the intervention within the context of a patient’s unique phenotypic profile.^33^ This can be achieved by defining biomarkers or parameters that describe mechanistic pathways through which an intervention exerts its beneficial effects,^34^ such as through molecular or proteomic profiling.^17,35^ Yet these mechanisms are not always known *a priori*, and molecular analyses are costly and, therefore, ineligible for use in large studies. N-of-1 trials, which involve periodic switching between treatment arms such that each individual functions as their own control have been proposed as a potential mechanism to personalize treatment effects.^36^ Despite their promise, this trial design is not applicable to large, phase III trials powered for major clinical endpoints. Similar concepts also apply to response adaptive randomization,^37^ or sequential multiple assignment adaptive randomized trials (SMART),^38^ which allow patients who do not respond to an initial treatment to be re-randomized.

In this context, our method bridges an adaptive trial design with predictive enrichment through machine learning-driven insights into phenotypes associated with individualized treatment effects. First, our method relies on an approach that can be defined *a priori* and remains independent of investigator input or biases for the duration of the study. Second, it does not require prior knowledge on potential phenotypic determinants of heterogeneous treatment effects but rather learns those in real-time. Third, the algorithm adaptively conditions the probability of enrollment on the predicted treatment benefit, thus enabling the trial to enrich its sample for phenotypes more likely to benefit. However, this process is not deterministic, and the algorithm incorporates a level of uncertainty which enables the continued recruitment of both high and low responders at different rates. As such, it does not change the target population that gets screened but may be used as a gatekeeper prior to randomization to maximize the efficient use of limited resources. Fourth, by modeling the relative treatment effect, our algorithm provides predictive enrichment without restricting enrollment to the individuals most likely to experience the primary endpoint. Depending on available resources, this could be used to selectively enrich the trial population for certain phenotypes without restricting access to any individuals. However, in the setting of limited resources, this can be used to prioritize phenotypes that demonstrate greater net benefit. Fifth, across simulations where there is an absence of heterogeneity in the treatment effect, the model does not result in net predictive enrichment. Finally, it respects the random treatment assignment throughout the trial since the predictive algorithm is trained on information collected before randomization.

Given the experimental approach, clinical translation of this method would require several key assumptions to be made *a priori*, which depend on the intervention and study population. In fact, there is precedent for clinical decision-making driven by post-hoc analyses of trials showing heterogeneous treatment effects.^34,39^ Depending on the available resources and risk of the intervention, one option may be to adapt the size of the trial while maximizing efforts to enroll the highest responders without restricting access to any patients. Alternatively, investigators may choose to embed a risk stratification algorithm that can direct a study of a high-risk intervention with limited resources towards participants more likely to derive net benefit. On this note, the implications for the regulatory submission as well as feedback from individual stakeholders (sponsor, investigators, patient representatives) may determine the optimal approach, which might differ between therapeutic interventions (medications or devices) and screening or diagnostic strategies. Finally, the estimand needs to be individualized for each study to best estimate the treatment effect while accounting for possible intercurrent events. An example is deciding whether intention-to-treat (ITT) or as-protocol analyses should be used to define individualized effect estimates using the phenomapping method.^40,41^

Our analysis carries limitations that merit consideration. First, our work here represents a post hoc analysis of real-world RCTs. However, IRIS was chosen given that it illustrates frequent challenges faced in cardiovascular outcome trials, including but not limited to slow enrollment (∼7-8 years), the need for a large study group (∼almost 4000 patients), and long prospective follow-up (∼5 years). Similarly, SPRINT models a large cardiovascular trial of a strategy rather than a specific medication. Second, the regulatory implications of new adaptive designs are not well-defined, such as determining the product label for therapies approved based on similar trials. It should be noted, however, that our proposed design does not explicitly incorporate new exclusion or inclusion criteria but rather modifies the probability that a patient fulfilling all original inclusion criteria will be enrolled at the later stages of the trial. Notably, post hoc analyses of completed trials have demonstrated variation across sites in the characteristics as well as outcomes of the enrolled populations, highlighting existing variations despite specific protocols and enrollment criteria.^42^ Third, a predictively enriched, accelerated study design could hinder our ability to identify safety signals for rare events or derive inference from traditional subgroup analyses, including populations with traditionally low rates of events. Fourth, we cannot disentangle the effects of deploying a model at interim timepoints from the effects of phenomapping specifically. However, our prior work has demonstrated the ability of our approach to detect modest embedded heterogeneous treatment effects in an interpretable manner, and our inability to dissect the source of benefit is not expected to have practical implications in deployment.^11^ Finally, although the algorithm aims to introduce explainability to its predictions through SHAP, reliance on broad phenotypic features is often a surrogate for biological, functional, or anatomical differences. Similarly, the patterns for enrichment are specific to each trial and do not allow generalizable conclusions on how the enriched population may look for a different trial. Nevertheless, the algorithm can be tuned as appropriate to be more or less conservative in its predictions and recommendations. It can also incorporate high-dimensional features, such as genomic, or imaging biomarkers.

In conclusion, we describe and implement an ML-guided algorithm for adaptive, predictive enrichment of RCTs based on individualized signatures of treatment benefit derived from computational trial phenomaps. In a post hoc analysis of two large cardiovascular outcome trials powered for clinical endpoints as primary outcomes, our proposed strategy of predictive enrichment based on ML-derived insights achieves a consistent and robust reduction in the required sample size while conserving the study’s power to detect significant average treatment effects. This is achieved through real-time predictive enrichment which is independent of a patient’s baseline absolute risk and modifies the trial’s baseline phenotypic composition in a standardized way, thus promoting a trial’s efficiency, safety, power, and generalizability.

## METHODS

### Data source and patient population

The Insulin Resistance Intervention after Stroke (IRIS) trial recruited patients at least 40 years of age with a recent ischemic stroke or a transient ischemic attack (TIA) during the 6 months prior to randomization, who did not have diabetes mellitus at the time of enrollment but had evidence of insulin resistance based on a homeostasis model assessment of insulin resistance (HOMA-IR) index score of 3.0 or greater. Participants were randomly assigned in a 1:1 ratio to receive either pioglitazone or matching placebo (with dose up-titration as specified in the original trial report).^18^ Patients were contacted every 4 months, and participation ended at 5 years or at the last scheduled contact before July 2015. The Systolic Blood Pressure Intervention Trial (SPRINT) enrolled 9361 participants, 50 years of age or older, with a systolic blood pressure (SBP) of 130-180 mm Hg with or without antihypertensive drug treatment as well as an additional indicator of cardiovascular risk. These included clinical or subclinical cardiovascular disease, chronic kidney disease, 10-year risk Framingham Risk Score of cardiovascular disease of 15% or higher or age of 75 years or older. Patients with diabetes mellitus, prior stroke, or dementia were excluded from this trial. Participants were enrolled between 2010-2013 at 102 clinical sites in the U.S.^19^

### Study outcomes

In accordance with the primary outcome of the original trials, we focused on a composite of first fatal or nonfatal stroke or fatal or nonfatal myocardial infarction as the primary outcome for IRIS, and a composite of myocardial infarction, acute coronary syndrome not resulting in myocardial infarction, stroke, acute decompensated heart failure, or death from cardiovascular causes for SPRINT. We also explored a hierarchical composite outcome as follows; IRIS: all-cause mortality, followed by non-fatal MACE (major adverse cardiovascular events) components, hospitalization events, heart failure events, and bone fractures; SPRINT: all-cause mortality, followed by non-fatal MACE and finally serious adverse events. Definitions were concordant with those used in the original trial reports.^18,19^ All outcomes and selected safety events were adjudicated by the members of independent committees in a blinded fashion for each of the trials.

### Design of a group sequential, adaptive trial experiment

We designed a simulation algorithm to test the hypothesis that interim ML-guided analyses of computational trial phenomaps can adaptively guide the trials’ enrollment process and maximize their efficiency while reducing their final/required size. The tenet of this approach is that ML phenomapping-derived insights can steer the recruitment towards patients who are more likely to benefit from the intervention. For this, we defined three interim analysis timepoints, with the final analysis occurring once all primary events had been reported. It should be noted that the original power calculation for IRIS had assumed higher event and faster enrollment rates than the ones that were observed during the trial, thus prompting serial amendments to the trial protocol, including an extension of recruitment and an increase in the study size (from 3136 patients initially to 3936 patients). In a post-hoc fashion, knowing that the primary outcome occurred in 228 of 1937 participants in the placebo arm (∼11.8% rate) and 175 of 1939 participants in the pioglitazone arm (∼9.0%), we simulated power calculations assuming a superiority trial design with a one-sided α of 0.025 (see “*power calculations*” below). We defined the timepoint at which 50, 100 and 150 total primary outcome events had been recorded in the original trial as the timepoint for our first, second and third interim analysis timepoints, respectively. In SPRINT we assumed that the respective primary outcome would occur in 6.8% vs 5.4% of the standard and intensive arms, and for consistency, defined the interim analysis timepoints based on the occurrence of the first 50, 100 and 150 primary outcome events.

### Overview of the predictive enrichment approach

During the first enrollment period of the simulation (study onset until first interim analysis) we enrolled all trial participants, similar to the original trials, without any restrictions or modifications in the enrollment process. Beginning at the first interim analysis timepoint, participants recruited up until that stage were randomly split into training/cross-validation (50%) and testing sets (50%). In the training set, baseline data were pre-processed and used to define a phenomap (see “*Machine learning trial phenomapping*” below), which represented the phenotypic architecture of the population across all axes of baseline variance. Through iterative analyses centered around each unique individual and weighted for each individual participant’s location in the phenotypic space, we defined individualized estimates of the effects of the studied intervention, as compared to the control arm, for the primary outcome.

Subsequently, we built a ML framework to identify key features that collectively determined a phenotypic signature (algorithm) predictive of these individualized estimates. The algorithm was then applied in the testing set, assessing for evidence of potential heterogeneous treatment effects by dichotomizing the population into two groups based on their predicted response. To avoid imbalanced groups or extreme outliers of responders or non-responders, the smallest subgroup size was set at 20%. We then analyzed the presence of heterogeneity in the observed effect estimates between the two subgroups in the testing set by calculating the *p* value for interaction of treatment effect. Given that testing was done using just a half of the observations collected at each interim analysis timepoint, we defined a threshold of *p_interaction_*<0.2 as our criterion for possible presence of heterogeneity.

If there was potential evidence of heterogeneity based on this analysis, sample size calculations were updated at that interim analysis timepoint by revising the expected effect size (under the assumption of predictive enrichment) at the original power and alpha levels (0.8 and 0.025, respectively, in both trials). This was done to assess whether prospective predictive enrichment and the associated decrease in the projected number of recruited individuals would provide sufficient power at a sample size equal to or smaller than the originally planned one. We performed sample size calculations assuming various levels of predictive enrichment, which ranged from enrolling 50% to 95% of all remaining candidates, in 5% increments. If there were several levels that met these criteria, we ultimately chose the predictive enrichment level that minimized the required sample.

Assuming the above, over the subsequent period, the probability of enrollment was conditioned on the anticipated benefit, assessed by applying the most recent model to each potential candidate’s baseline characteristics. Alternatively, if there was no evidence of heterogeneous treatment effect, or the proposed enrichment in enrollment would not be adequately powered at the revised sample size, we proceeded as originally planned and continued with standard enrollment for that time-period without predictive enrichment. This process was repeated at each interim analysis timepoint. There was no assessment for futility.

Given the stochastic nature of the algorithm, the simulation was repeated *r*=10 times. For reference, we present the observed outcomes of the full trial population at the same timepoints. To enable direct comparison between the different simulations, the final analysis was performed at the timepoint at which all primary outcome events had occurred in the original trial population.

### Data-preprocessing

Our analysis included 62 phenotypic features recorded at baseline in IRIS (**Supplementary Table 2**), and 82 baseline features in SPRINT (**Supplementary Table 3**), as per our prior work.^11^ At every point, pre-processing steps, including imputation, were performed independently for each patient subset to avoid data leakage. Baseline features with greater than 10% missingness were removed from further analysis. To avoid collinearity of continuous variables, we calculated pairwise correlations across variables, and wherever pairs exceeded an absolute correlation coefficient of 0.9, we excluded the variable with the largest mean absolute correlation across all pairwise comparisons. Continuous variables also underwent 95% winsorization to reduce the effects of extreme outliers, whereas factor variables with zero variance were dropped from further processing. Next, we imputed missing data using a version of the random forest imputation algorithm adapted for mixed datasets with a maximum of five iterations (function MissForest, from Python package missingpy v0.2.0). Factor variables underwent one-hot encoding for ease of processing with downstream visualization and machine learning algorithms.

### Computational trial phenomaps

Once the dataset for a given simulation at a specified time-point was created, we computed a dissimilarity index that classified individuals based on their detailed clinical characteristics according to Gower’s distance. Gower’s method computes a distance value for each pair of individuals. For continuous variables, Gower’s distance represents the absolute value of the difference between a pair of individuals divided by the range across all individuals. For categorical variables, the method assigns “1” if the values are identical and “0” if they are not. Gower’s distance is ultimately calculated as the mean of these terms.^43^ At this point, the phenotypic architecture of the trial can be visualized using uniform manifold approximation and projection (UMAP),^44^ a method that constructs a high-dimensional graph and then optimizes a low-dimensional graph to be as structurally similar as possible.

UMAP aims to maintain a balance between the local and global structure of the data by decreasing the likelihood of connection as the outwards radius around each data point increases, thus maintaining the local architecture while ensuring that each point is connected to at least its closest neighbor and ensuring a global representation.^44^

### Defining individualized hazard estimates

To extract personalized estimates of predicted benefit with pioglitazone or intensive SBP control, versus placebo or standard SBP reduction respectively, for each individual included in each interim analysis, we applied weighted estimation in Cox regression models.^45^ We used all pertinent outcome events for the selected population (in the training set) with censoring at the time of the specified interim analysis. With every iteration of this regression around each unique individual, every study participant was assigned unique weights based on the phenotypic (Gower’s) distance from the index patient of that analysis. To ensure that patients phenotypically closer to the index patient carried higher cumulative weights than patients located further away, we applied a cubic exponential transformation of the similarity metric, defined as (*1-Gower’s distanc*e). These values were further processed through a Rectified Linear Unit (ReLU) function prior to their inclusion as weights in the regression models. This allowed us to simultaneously model an exponential decay function and control the impact of low values (ReLU). From each personalized Cox regression model (fitted for each unique participant with individualized weightings as above), we extracted the natural logarithmic transformation of the hazard ratio (log HR) for the primary outcome for the intervention versus control.

### Machine learning estimation of individualized benefit

To identify baseline features that are important in determining the personalized benefit of the studied intervention relative to control (described by the individualized log HR), an extreme gradient boosting algorithm (known as XGBoost; based on a tree gradient booster) was fitted with simultaneous feature selection based on the Boruta and SHAP (SHapley Additive exPlanations) methods. Briefly, the Boruta method creates randomized (permuted) versions of each feature (called “shadow features”) before merging the permuted and original data. Once a model is trained, the importance of all original features is compared to the highest feature importance of the shadow features. This process was repeated for n=20 iterations, without sampling. SHAP was added as an approach to explain the output of the ML model, based on concepts derived from game theory. SHAP calculates the average marginal contributions for each feature across all permutations at a local level. With the addition of SHAP analysis, the feature selection further benefits from the strong additive feature explanations but maintains the robustness of the Boruta method.^46^ The testing data were further split into training and testing sets (with a random 80-20% split). We set our problem as a regression task using root mean squared error as our metric to evaluate our model’s accuracy during testing. Before training, the labels (i.e., previously calculated individualized log HR) underwent 95% winsorization to minimize the effects of extreme outliers. First, we fitted an XGBoost model using the Boruta algorithm to identify a subset of important baseline features, and then repeated this process to predict the individualized log HR, this time using only the selected features as input. Hyperparameter tuning was achieved through a grid search across 25 iterations (learning rate: [0.01, 0.05, 0.10, 0.15]; maximal depth of the tree: [3, 5, 6, 10, 15, 20]; fraction of training samples used to train each tree: 0.5 to 1.0 by 0.1 increments, number of features supplied to a tree: 0.4 to 1.0 by 0.1 increments; random subsample of columns when every new level is reached: 0.4 to 1.0 by 0.1, number of gradient boosted trees: [100, 500, 1000]). We trained the model for a maximum of 1000 rounds, with an early stopping function every 20 rounds once the loss in the validation set started to increase. The importance of each feature was again visualized using a SHAP plot. SHAP values measure the impact of each variable considering the interaction with other variables. We visualized these using a SHAP summary plot, in which the vertical axis represented the variables in descending order of importance and the horizontal axis indicated the change in prediction. The gradient color denotes the original value for that variable (for instance, for binary variables such as sex, it only takes two colors, whereas for continuous variables, it contains the whole spectrum). In the **Supplementary Table 6** we present a more extensive discussion of key parameters and the rationale behind their default values.

### Adaptive trial enrollment

Once a predictive model was generated at a given interim analysis timepoint, the model was prospectively applied to all trial candidates during the following trial period (time between two interim analyses). For example, a model trained at interim analysis timepoint #1 was applied to individuals screened between the interim analysis timepoints #1 and #2. Here, all patients that were included and enrolled in the original trial during this period were considered eligible candidates. This approach yielded individualized predictions of expected cardiovascular benefit with pioglitazone versus placebo (or intensive versus standard SBP reduction), with these predictions used to condition the probability of a given patient being enrolled in the simulation. Given that the predicted individualized log HR could have both negative (favoring pioglitazone or intensive SBP reduction) and positive values (favoring placebo or standard SBP reduction), predictions were multiplied by –1, normalized to the [0, 1] range. The result (input *x*) was processed through a sigmoid transformation function (**Equation (1)**) with a scaling factor of *k*=10; where *z* = the ratio of the responders to non-responders, followed by squared transformation.

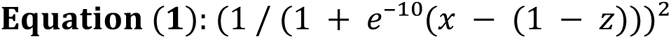

These numbers were used as sampling weights during the subsequent period to ensure that patients with higher predicted benefit were more likely to be enrolled over the next period. The process was repeated at each interim analysis timepoint.

### Power calculations

We simulated a superiority trial design, assuming a power of 80% and type I error of 0.025. We present alpha level adjustments for each time point, adjusted based on the O’Brien-Fleming and alpha-spending Pocock methods (**Supplementary Table 1**). We performed our analyses using the *rpact* package in R, using the expected event rates used in our power calculations above and simulating three interim analyses with four total looks. These analyses are restricted to the primary endpoint.

### Negative control analysis

To assess the performance of our algorithm in the presence of an identical average treatment effect (ATE) but with absent (or at least randomly distributed) heterogeneous treatment effects, we randomly shuffled the baseline characteristics of each trial. This ensured that any effects of the baseline characteristics on the effectiveness of the intervention would be lost or be due to random variation.

### Stability analysis

To assess the robustness of our estimates, we defined three interim analysis timepoints for each study. At each timepoint, we randomly split the respective study population into a training and testing set (50:50 split) and repeated the process 100 times (using 100 random seeds). Across these 100 iterations, each observation (study participant) was roughly represented in the training set in half of all splits, and in the testing set in the remaining half. This enabled the variation of the demographic and baseline characteristics of each training population. For each iteration, we followed the same phenomapping approach presented in our work, estimated individualized hazard ratios, fit an extreme gradient boosting (XGBoost) regressor, and subsequently used this to infer personalized effect estimates in the testing set. As done in our main analysis, individuals in the testing set were then ranked based on their predicted responses. Using this approach, we defined the following stability metrics:

*Concordance of relative ranks based on XGBoost-defined personalized treatment effects*: a metric reflecting the stability of relative ranks for two random observations/study participants. For a random patient A and patient B, this reflects the probability that patient A will be consistently ranked as a higher responder than patient B (or vice versa) across simulations, when both patients A and B are present in the testing set. A value of 1 (number of times A>B is equal to the times B>A) indicates that there is no consistent pattern in the relative ranks, whereas higher values suggest greater concordance in the relative ranks. For this, A generally refers to the patient who is most often listed as a higher responder than patient B. This allows the calculation of a concordance metric (C(A,B)) for each pair of individuals (**Equation (2)**). The mean concordance across all possible pairs of individuals is then calculated based on **Equation (3)** which estimates the average of the upper triangle of the concordance matrix (as further described in **Supplementary** Figure 1). Using this concept, a concordance ratio can then be computed based on the number of times there was concordance vs discordance in the relative ranks across all observations. 95% Confidence intervals are calculated using the bootstrapping method with n=1000 replications.

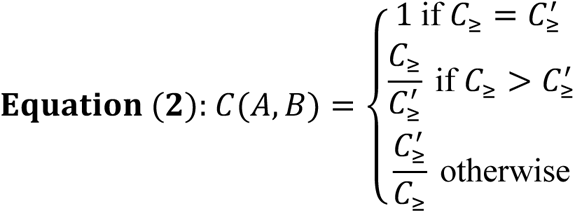

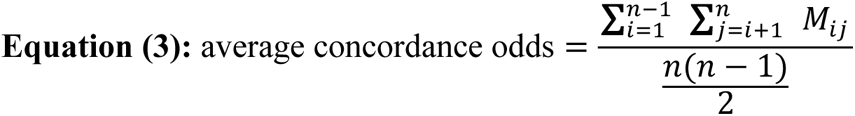

*Consistency of Boruta SHAP-defined features*: Along with the concordance of relative prediction ranks, we also record the features selected by the Boruta SHAP algorithm at each interim analysis timepoint, and then present the relative frequency with which a given feature presents across iterations and timepoints. This approach assesses the general consistency in the features identified as potential predictors of heterogeneous treatment effects across distinct time periods. It should be noted that features are not independent of each other, and therefore the algorithm may detect different features during each iteration that collectively define a similar patient phenotype.

### Statistical analysis

Categorical variables are summarized as numbers (percentages) and continuous variables as mean ± standard deviation (unless specified otherwise) or median with IQR (Q1 to Q3) unless specified otherwise. Categorical groupings (i.e., study arm assignment) across adaptive runs and the original trial were compared using the chi-square test. Survival analyses were performed by fitting a Cox regression model for the time-to-primary outcome using the treatment arm as an independent predictor. While estimating individualized treatment effects, each observation was weighted based on the calculated similarity metric to the index patient of each analysis. Between-subgroup analyses for heterogeneity of treatment effect were performed by computing a *p* value for interaction. Simulation-level counts and point estimates were compared to the respective numbers/counts from the original trial using one-sample t-tests; for the counts of final study participants, alpha was set at 0.025 given the single-sided nature of the test; otherwise, alpha was set at 0.05. We graphically summarized the counts of enrolled participants, primary outcome events, Cox regression-derived effect estimates (unadjusted, for each one of the primary and secondary outcomes), and *p* values at each one of the interim timepoints (with error bars denoting the standard error of the mean), in addition to the final analysis timepoint. Cumulative incidence curves for the primary outcome stratified by the enrollment period and simulation analysis were graphically presented. Each one was compared to the original trial subset for the same period using the log-rank statistic. An exploratory hierarchical outcome analysis was assessed using the win ratio for prioritized outcomes and corresponding 95% confidence intervals.^47^ As reviewed above, for the primary outcome, we simulated a superiority trial design, assuming a power of 80% and type I error of 0.025. Statistical tests were two-sided with a level of significance of 0.05, unless specified otherwise. Analyses were performed using Python (version 3.9) and R (version 4.2.3), and reported according to STROBE guidelines.^48^ Package used for analysis are specified in the **Supplementary Methods**.

### Ethics approval

The local ethics committee/IRB (institutional review boards) of Yale University provided a determination of exemption and waived ethical approval for this work (under IRB protocol ID 2000029730, 1/21/2021).

## DATA AVAILABILITY

The SPRINT data are available through the Biologic Specimen and Data Repository Information Coordinating Center (BioLINCC) of the National Heart, Lung, and Blood Institute (NHLBI). The IRIS data were made available through communication with the original study investigators.

## CODE AVAILABILITY

The code for the study is available from the authors upon request.

## ACKNOWLEDGEMENTS

The authors acknowledge funding through the Yale-Mayo Center of Excellence in Regulatory Science and Innovation (CERSI) (EKO), the National Heart, Lung, and Blood Institute (NHLBI) of the National Institutes of Health (NIH) under awards 1F32HL170592-01 (EKO) and K23HL153775 (RK), and the Doris Duke Charitable Foundation under award 2022060 (RK). The content is solely the responsibility of the authors and does not necessarily represent the official views of the funders. The authors would like to thank Walter N. Kernan (Department of Internal Medicine, Yale School of Medicine) for facilitating access to the IRIS data.

## AUTHOR CONTRIBUTIONS

R.K. and E.K.O. conceived the study and assessed the data. E.K.O. pursued the analytical work, wrote the code for the analysis, and drafted the manuscript. All authors provided feedback regarding the study design, made critical contributions to writing of the manuscript, and approved its final draft. R.K. supervised the study, procured funding. E.K.O. and R.K. are responsible for the accuracy of the analysis.

## COMPETING INTERESTS

E.K.O. declares funding through the National Heart, Lung, and Blood Institute of the National Institutes of Health (1F32HL170592-01), is a co-inventor of the U.S. Patent Applications 63/508,315 & 63/177,117, and has previously served as a consultant to Caristo Diagnostics Ltd (outside the present work). R.K. is an Associate Editor of JAMA. He receives support from the National Heart, Lung, and Blood Institute of the National Institutes of Health (under award K23HL153775) and the Doris Duke Charitable Foundation (under award, 2022060). He also receives research support, through Yale, from Bristol-Myers Squibb and Novo Nordisk. He is a coinventor of U.S. Provisional Patent Applications 63/177,117, 63/428,569, 63/346,610, 63/484,426, and 63/508,315. E.K.O. and R.K. are co-founders of Evidence2Health, a precision health platform to improve evidence-based cardiovascular care. D.L.B. discloses the following relationships – Advisory Board: Angiowave, Bayer, Boehringer Ingelheim, Cardax, CellProthera, Cereno Scientific, Elsevier Practice Update Cardiology, High Enroll, Janssen, Level Ex, McKinsey, Medscape Cardiology, Merck, MyoKardia, NirvaMed, Novo Nordisk, PhaseBio, PLx Pharma, Regado Biosciences, Stasys; Board of Directors: Angiowave (stock options), Boston VA Research Institute, Bristol Myers Squibb (stock), DRS.LINQ (stock options), High Enroll (stock), Society of Cardiovascular Patient Care, TobeSoft; Chair: Inaugural Chair, American Heart Association Quality Oversight Committee; Consultant: Broadview Ventures, Hims; Data Monitoring Committees: Acesion Pharma, Assistance Publique-Hôpitaux de Paris, Baim Institute for Clinical Research (formerly Harvard Clinical Research Institute, for the PORTICO trial, funded by St. Jude Medical, now Abbott), Boston Scientific (Chair, PEITHO trial), Cleveland Clinic (including for the ExCEED trial, funded by Edwards), Contego Medical (Chair, PERFORMANCE 2), Duke Clinical Research Institute, Mayo Clinic, Mount Sinai School of Medicine (for the ENVISAGE trial, funded by Daiichi Sankyo; for the ABILITY-DM trial, funded by Concept Medical), Novartis, Population Health Research Institute; Rutgers University (for the NIH-funded MINT Trial); Honoraria: American College of Cardiology (Senior Associate Editor, Clinical Trials and News, ACC.org; Chair, ACC Accreditation Oversight Committee), Arnold and Porter law firm (work related to Sanofi/Bristol-Myers Squibb clopidogrel litigation), Baim Institute for Clinical Research (formerly Harvard Clinical Research Institute; RE-DUAL PCI clinical trial steering committee funded by Boehringer Ingelheim; AEGIS-II executive committee funded by CSL Behring), Belvoir Publications (Editor in Chief, Harvard Heart Letter), Canadian Medical and Surgical Knowledge Translation Research Group (clinical trial steering committees), CSL Behring (AHA lecture), Cowen and Company, Duke Clinical Research Institute (clinical trial steering committees, including for the PRONOUNCE trial, funded by Ferring Pharmaceuticals), HMP Global (Editor in Chief, Journal of Invasive Cardiology), Journal of the American College of Cardiology (Guest Editor; Associate Editor), K2P (Co-Chair, interdisciplinary curriculum), Level Ex, Medtelligence/ReachMD (CME steering committees), MJH Life Sciences, Oakstone CME (Course Director, Comprehensive Review of Interventional Cardiology), Piper Sandler, Population Health Research Institute (for the COMPASS operations committee, publications committee, steering committee, and USA national co-leader, funded by Bayer), Slack Publications (Chief Medical Editor, Cardiology Today’s Intervention), Society of Cardiovascular Patient Care (Secretary/Treasurer), WebMD (CME steering committees), Wiley (steering committee); Other: Clinical Cardiology (Deputy Editor), NCDR-ACTION Registry Steering Committee (Chair), VA CART Research and Publications Committee (Chair); Patent: Sotagliflozin (named on a patent for sotagliflozin assigned to Brigham and Women’s Hospital who assigned to Lexicon; neither I nor Brigham and Women’s Hospital receive any income from this patent); Research Funding: Abbott, Acesion Pharma, Afimmune, Aker Biomarine, Alnylam, Amarin, Amgen, AstraZeneca, Bayer, Beren, Boehringer Ingelheim, Boston Scientific, Bristol-Myers Squibb, Cardax, CellProthera, Cereno Scientific, Chiesi, CinCor, Cleerly, CSL Behring, Eisai, Ethicon, Faraday Pharmaceuticals, Ferring Pharmaceuticals, Forest Laboratories, Fractyl, Garmin, HLS Therapeutics, Idorsia, Ironwood, Ischemix, Janssen, Javelin, Lexicon, Lilly, Medtronic, Merck, Moderna, MyoKardia, NirvaMed, Novartis, Novo Nordisk, Otsuka, Owkin, Pfizer, PhaseBio, PLx Pharma, Recardio, Regeneron, Reid Hoffman Foundation, Roche, Sanofi, Stasys, Synaptic, The Medicines Company, Youngene, 89Bio; Royalties: Elsevier (Editor, Braunwald’s Heart Disease); Site Co-Investigator: Abbott, Biotronik, Boston Scientific, CSI, Endotronix, St. Jude Medical (now Abbott), Philips, SpectraWAVE, Svelte, Vascular Solutions; Trustee: American College of Cardiology; Unfunded Research: FlowCo, Takeda. H.M.K. works under contract with the Centers for Medicare & Medicaid Services to support quality measurement programs, was a recipient of a research grant from Johnson & Johnson, through Yale University, to support clinical trial data sharing; was a recipient of a research agreement, through Yale University, from the Shenzhen Center for Health Information for work to advance intelligent disease prevention and health promotion; collaborates with the National Center for Cardiovascular Diseases in Beijing; receives payment from the Arnold & Porter Law Firm for work related to the Sanofi clopidogrel litigation, from the Martin Baughman Law Firm for work related to the Cook Celect IVC filter litigation, and from the Siegfried and Jensen Law Firm for work related to Vioxx litigation; chairs a Cardiac Scientific Advisory Board for UnitedHealth; was a member of the IBM Watson Health Life Sciences Board; is a member of the Advisory Board for Element Science, the Advisory Board for Facebook, and the Physician Advisory Board for Aetna; and is the co-founder of Hugo Health, a personal health information platform, and co-founder of Refactor Health, a healthcare AI-augmented data management company. M.A.S. reports institutional grant support from the US National Institutes of Health, US Food and Drug Administration, and US Department of Veteran Affairs; personal consulting fees from Janssen Research and Development and Private Health Management; and institutional grant support from Advanced Micro Devices, outside the scope of the submitted work. All other authors declare no competing interests.

## Notes

### Summary of Updates

We have implemented additional stability analyses suggested by peer reviewers, expanded the number of simulations runs, and re-written parts of the introduction and discussion. As a result, the figures and corresponding data, results, and supplement have all been revised.

