## Supplemental Material for "An explainable machine learning-based phenomapping strategy for adaptive predictive enrichment in randomized controlled trials"

### Supplementary Information

#### Supplementary Methods

**Statistical and programming packages used:**

Python 3.9:
- numpy=1.23.5
- pandas=1.5.3
- python=3.9.16
- autograd==1.5
- borutashap==1.0.16
- gower==0.0.5
- hyperopt==0.2.7
- lifelines==0.26.4
- matplotlib==3.3.3
- missingpy==0.2.0
- scikit-learn==0.24.0
- scipy==1.10.1
- seaborn==0.11.2
- shap==0.39.0
- statsmodels==0.13.2
- tabula==1.0.5
- xgboost==1.6.2

R 4.2.3
- WinRatio=1.0
- powerSurvEpi=0.1.3
- survival=3.5.5
- Publish=2023.1.17
- purr=1.0.1
- dplyr=1.1.2

#### Supplementary Tables

**Supplementary Table 1. Power calculations and alpha spending adjustments for group sequential simulations.**

| | Interim analyses: | | | Final analysis |
|---|---|---|---|---|
| | **#1 (50 events)** | **#2 (100 events)** | **#3 (150 events)** | |
| **IRIS** | | | | |
| O'Brien-Fleming | 5.58, <0.0001 | 3.93, <0.0001 | 3.209, 0.0007 | 1.965, 0.0250 |
| Pocock | 2.402, 0.0082 | 2.402, 0.0142 | 2.402, 0.0188 | 2.402, 0.0250 |
| Alpha-spending Pocock | 2.585, 0.0049 | 2.558, 0.0089 | 2.529, 0.0124 | 2.173, 0.0250 |
| **SPRINT** | | | | |
| O'Brien-Fleming | 6.514, <0.0001 | 4.604, 0.0003 | 3.761, 0.0108 | 1.961, 0.0250 |
| Pocock | 2.410, 0.0080 | 2.410, 0.0139 | 2.410, 0.0184 | 2.410, 0.0250 |
| Alpha-spending Pocock | 2.686, 0.0036 | 2.652, 0.0068 | 2.617, 0.0096 | 2.121, 0.0250 |

Assumptions: alpha=0.025 (one-sided), beta=0.80. Values presented as efficacy boundary (z-value scale), cumulative alpha spent. IRIS: Insulin Resistance Intervention after Stroke; SPRINT: Systolic Blood Pressure Intervention Trial.

**Supplementary Table 2. Summary of baseline features included in IRIS.**

| | |
|---|---|
| Age at time of blood test<br>Modified Rankin grade<br>Gender<br>Ethnicity<br>Race<br>Waist-to-hip ratio<br>Body mass index<br><br>Prior history of stroke or transient ischemic attack<br>History of diabetes<br>Oral steroid use<br>History of cancer<br>History of liver disease<br>History of lung disease<br>History of kidney disease<br>History of congestive heart failure<br>NYHA class<br>Carotid artery stenosis<br>Atrial fibrillation<br>Myocardial infarction history<br>History of coronary artery bypass grafting surgery<br>History of coronary angioplasty<br>History of hypertension<br>History of peripheral vascular disease<br>History of hypercholesterolemia<br>Smoking: ever<br>Smoking: current<br>Current alcohol use<br>Lower extremity edema<br><br>Ejection fraction (%)<br><br>Systolic blood pressure<br>Diastolic blood pressure | Medications:<br>- Oral hypoglycemics<br>- Thiazolidinediones<br>- Insulin use<br>- Non-insulin injectable agents<br>- Oral steroids<br>- ACEi (angiotensin-converting enzyme inhibitor) or ARB (angiotensin II-receptor blocker)<br>- Beta-blockers<br>- Calcium-channel blockers<br>- Thiazide diuretics<br>- Loop diuretics<br>- Statins<br>- Fibrates<br>- Antiarrhythmics<br>- Anticoagulants<br>- Antidepressants<br>- Sedatives and hypnotics<br>- Hormone replacement<br>- Antiplatelet therapy<br>- Mineralocorticoid antagonists<br>- Potassium-sparing diuretics<br>- Use of any antihypertensive therapy<br><br>Laboratory measurements:<br>- Fasting glucose<br>- HOMA: *Homeostatic Model Assessment for Insulin Resistance*<br>- Total cholesterol<br>- High-density lipoprotein (HDL) levels<br>- Low-density lipoprotein (LDL) levels<br>- Triglycerides<br>- Alanine transaminase (ALT) levels<br>- Hemoglobin<br>- C-reactive protein<br>- Hemoglobin A1c<br>- Bone density measure |

IRIS: Insulin Resistance Intervention after Stroke.

**Supplementary Table 3. Summary of baseline features included in SPRINT.**

| | |
|---|---|
| Age | Patient-reported history of heart attack |
| Female sex | Patient-reported history of irregular heartbeat |
| Race (White, Black, Hispanic, Other) | Patient-reported history of osteoarthritis |
| Smoking | Patient-reported history of rheumatoid arthritis |
| History of cardiovascular disease | Patient-reported history of gout |
| History of chronic kidney disease | Patient-reported history of other arthritis |
| History of myocardial infarction | Patient-reported history of hip problems |
| History of coronary revascularization | Patient-reported history of cancer |
| Atrial fibrillation/flutter | Patient-reported history of skin cancer |
| History of angina | Patient-reported history of peripheral vascular |
| Family history of cardiovascular disease | disease |
| Family history of premature cardiovascular disease | Patient-reported history of thyroid disease |
| Days per week of vigorous activity | Patient-reported history of anemia |
| Patients reports fainting when standing | Patient-reported history of hypertension |
| Left ventricular hypertrophy by electrocardiography | Patient-reported history of low back pain |
| Body mass index | |
| | Use of blood pressure medications at baseline |
| Standing systolic blood pressure | Number of blood pressure medications |
| Standing diastolic blood pressure | Aspirin use |
| Standing heart rate | Statin use |
| Seating systolic blood pressure | |
| Seating diastolic blood pressure | Creatinine (serum) |
| Seating heart rate | Total cholesterol (serum) |
| Change in systolic blood pressure when standing | Blood urea nitrogen (serum) |
| Change in diastolic blood pressure when standing | Chloride (serum) |
| Change in heart rate when standing | Bicarbonate (serum) |
| | Creatinine (urine) |
| Montreal Cognitive Assessment (MOCA) score | Fasting glucose (serum) |
| Digit symbol substitution test score | High-density lipoprotein (serum) |
| | Potassium (serum) |
| Acute coronary syndrome | Low-density lipoprotein (serum) |
| Carotid disease | Sodium (serum) |
| Peripheral arterial disease revascularisation | Triglycerides (serum) |
| Known 50% or greater coronary, carotid, peripheral | Microalbumin (urine) |
| stenosis * | Urine microalbumin/creatinine ratio |
| Abdominal aortic aneurysm 5 cm or greater or repaired | Glomerular filtration rate |
| Calcium score 400 or greater | |
| Ankle-brachial index 0.9 or lower | QRS duration |
| Unable to stand | Cornell Voltage Product |
| Congestive heart failure | Cornell Voltage |
| Seizure | Sokolow-Lyon index |
| Stroke | Atrial fibrillation or flutter on electrocardiography |
| Transient ischemic attack | |
| Diabetes | Framingham risk score |
| Alcohol use disorder | Subjective assessment of general state of health |
| Dizziness | |

SPRINT: Systolic Blood Pressure Intervention Trial.

**Supplementary Table 4. Baseline characteristics for the final population across ten simulation runs in IRIS.**

| | Simulations | | | | | | | | | | Original |
|---|---|---|---|---|---|---|---|---|---|---|---|
| | #1 | #2 | #3 | #4 | #5 | #6 | #7 | #8 | #9 | #10 | |
| **Final number** | 2465 | 3512 | 3240 | 3298 | 3298 | 3512 | 2959 | 3802 | 3240 | 3709 | 3876 |
| **Pioglitazone arm** | 1236 (50.1) | 1759 (50.1) | 1631 (50.3) | 1655 (50.2) | 1642 (49.8) | 1773 (50.5) | 1478 (49.9) | 1901 (50.0) | 1623 (50.1) | 1852 (49.9) | 1939 (50.0) |
| **Age** | 63.0 (10.4) | 62.9 (10.6) | 63.1 (10.5) | 63.1 (10.5) | 63.1 (10.5) | 63.0 (10.6) | 62.5 (10.7) | 62.9 (10.6) | 62.8 (10.6) | 62.9 (10.6) | 62.9 (10.6) |
| **Female sex** | 877 (35.6) | 1203 (34.3) | 1160 (35.8) | 1178 (35.7) | 1186 (36.0) | 1113 (31.7) | 1028 (34.7) | 1308 (34.4) | 1107 (34.2) | 1280 (34.5) | 1338 (34.5) |
| **Asian race** | 27 (1.1) | 36 (1.0) | 33 (1.0) | 32 (1.0) | 31 (1.0) | 42 (1.2) | 32 (1.1) | 41 (1.1) | 35 (1.1) | 40 (1.1) | 42 (1.1) |
| **Black race** | 273 (11.3) | 374 (10.8) | 346 (10.9) | 335 (10.3) | 343 (10.6) | 394 (11.4) | 323 (11.1) | 403 (10.8) | 348 (10.9) | 404 (11.1) | 419 (11.0) |
| **White race** | 2086 (86.2) | 2988 (86.6) | 2759 (86.7) | 2826 (87.2) | 2819 (87.0) | 2960 (85.8) | 2512 (86.3) | 3237 (86.7) | 2749 (86.3) | 3146 (86.3) | 3291 (86.4) |
| **Hispanic ethnicity** | 91 (3.7) | 132 (3.8) | 117 (3.6) | 121 (3.7) | 122 (3.7) | 134 (3.8) | 115 (3.9) | 143 (3.8) | 120 (3.7) | 141 (3.8) | 147 (3.8) |
| **Prior history of stroke or TIA** | 379 (15.5) | 583 (16.7) | 555 (17.2) | 567 (17.3) | 589 (18.0) | 579 (16.6) | 450 (15.3) | 629 (16.6) | 590 (18.3) | 591 (16.0) | 643 (16.7) |
| **Hypertension** | 1840 (75.1) | 2433 (69.7) | 2328 (72.3) | 2367 (72.1) | 2376 (72.4) | 2497 (71.5) | 2054 (69.8) | 2713 (71.8) | 2226 (69.1) | 2638 (71.6) | 2770 (71.9) |
| **Atrial fibrillation history** | 167 (6.8) | 240 (6.9) | 227 (7.1) | 236 (7.2) | 239 (7.3) | 237 (6.8) | 169 (5.8) | 260 (6.9) | 223 (7.0) | 250 (6.8) | 264 (6.9) |
| **Myocardial infarction history** | 213 (8.7) | 300 (8.6) | 274 (8.5) | 285 (8.7) | 289 (8.8) | 296 (8.5) | 248 (8.4) | 320 (8.5) | 272 (8.4) | 310 (8.4) | 324 (8.4) |
| **Smoking (ever)** | 1657 (67.2) | 2338 (66.6) | 2196 (67.8) | 2200 (66.8) | 2199 (66.7) | 2339 (66.7) | 1963 (66.4) | 2522 (66.4) | 2160 (66.7) | 2464 (66.5) | 2564 (66.2) |
| **Hypercholesterolemia** | 1882 (77.2) | 2336 (67.2) | 2259 (70.4) | 2268 (69.5) | 2276 (69.7) | 2309 (66.4) | 2003 (68.4) | 2556 (67.9) | 2145 (66.9) | 2503 (68.2) | 2618 (68.2) |
| **Body mass index (kg/m²)** | 29.8 (5.5) | 29.8 (5.4) | 29.8 (5.4) | 29.8 (5.4) | 29.9 (5.4) | 29.7 (5.3) | 30.0 (5.5) | 29.9 (5.4) | 29.8 (5.4) | 29.9 (5.4) | 29.9 (5.4) |
| **Systolic blood pressure (mmHg)** | 133.3 (17.4) | 133.0 (17.2) | 133.1 (17.3) | 133.2 (17.5) | 133.3 (17.4) | 133.2 (17.4) | 132.9 (17.7) | 133.2 (17.5) | 133.1 (17.3) | 133.2 (17.6) | 133.2 (17.5) |
| **Diastolic blood pressure (mmHg)** | 79.1 (10.7) | 79.2 (10.6) | 79.2 (10.6) | 79.2 (10.7) | 79.2 (10.6) | 79.3 (10.6) | 79.1 (10.6) | 79.2 (10.6) | 79.3 (10.6) | 79.2 (10.6) | 79.2 (10.6) |
| **Modified Rankin scale** | 1.0 [0.0,2.0] | 1.0 [0.0,2.0] | 1.0 [0.0,2.0] | 1.0 [0.0,2.0] | 1.0 [0.0,2.0] | 1.0 [0.0,2.0] | 1.0 [0.0,2.0] | 1.0 [0.0,2.0] | 1.0 [0.0,2.0] | 1.0 [0.0,2.0] | 1.0 [0.0,2.0] |
| **HOMA** | 5.4 (2.9) | 5.5 (3.1) | 5.6 (3.1) | 5.5 (3.1) | 5.5 (3.1) | 5.5 (3.1) | 5.4 (2.9) | 5.5 (3.0) | 5.5 (3.1) | 5.5 (3.0) | 5.5 (3.0) |
| **LDL (mg/dL)** | 87.6 (31.8) | 87.2 (31.4) | 87.0 (31.4) | 86.7 (31.3) | 86.9 (31.4) | 86.9 (31.3) | 89.1 (32.0) | 87.7 (31.5) | 87.0 (31.3) | 87.8 (31.4) | 87.7 (31.5) |
| **HDL (mg/dL)** | 47.0 (12.9) | 47.3 (12.7) | 47.6 (12.9) | 47.5 (12.9) | 47.6 (12.8) | 47.1 (12.8) | 46.5 (12.6) | 47.1 (12.7) | 47.5 (12.8) | 47.0 (12.7) | 47.0 (12.7) |
| **Triglycerides (mg/dL)** | 142.7 (73.0) | 139.9 (72.3) | 139.8 (71.7) | 140.3 (73.0) | 140.0 (72.4) | 139.7 (73.0) | 143.3 (74.3) | 141.0 (73.0) | 139.6 (73.3) | 140.8 (72.5) | 141.0 (72.7) |
| **Antihypertensive therapy** | 2040 (82.8) | 2736 (77.9) | 2579 (79.6) | 2652 (80.4) | 2657 (80.6) | 2784 (79.3) | 2277 (77.0) | 3016 (79.3) | 2505 (77.3) | 2949 (79.5) | 3080 (79.5) |
| **Statin therapy** | 2062 (84.0) | 2888 (82.5) | 2701 (83.7) | 2753 (83.8) | 2738 (83.3) | 2894 (82.7) | 2395 (81.2) | 3123 (82.4) | 2659 (82.4) | 3035 (82.1) | 3186 (82.5) |
| **Antiplatelet agents** | 2274 (92.3) | 3220 (91.7) | 2976 (91.9) | 3028 (91.9) | 3023 (91.7) | 3219 (91.7) | 2731 (92.3) | 3489 (91.8) | 2965 (91.6) | 3411 (92.0) | 3557 (91.8) |
| **Anticoagulation** | 272 (11.1) | 398 (11.4) | 361 (11.2) | 375 (11.4) | 378 (11.5) | 399 (11.4) | 312 (10.6) | 433 (11.4) | 369 (11.4) | 418 (11.3) | 441 (11.4) |
| **ACEi or ARB** | 1529 (62.1) | 1857 (52.9) | 1837 (56.7) | 1908 (57.9) | 1894 (57.5) | 1957 (55.8) | 1693 (57.2) | 2117 (55.7) | 1764 (54.5) | 2057 (55.5) | 2154 (55.6) |
| **Beta-blockers** | 772 (31.4) | 1095 (31.3) | 991 (30.7) | 1047 (31.9) | 1053 (32.0) | 1094 (31.3) | 858 (29.1) | 1205 (31.8) | 925 (28.7) | 1182 (32.0) | 1228 (31.8) |

ACEi: Angiotensin-converting enzyme inhibitors; ARB: Angiotensin receptor blockers; HDL: High-density lipoprotein; HR: Hazard Ratio; HOMA: Homeostatic Model Assessment for Insulin Resistance; LDL: Low-density lipoprotein; TIA: transient ischemic attack. Continuous variables are summarized as mean (standard deviation) or median [25th-75th percentile], categorical variables as counts (valid percentages). All variables presented had <10% missing data at baseline.

**Supplementary Table 5. Baseline characteristics for the final population across ten simulation runs in SPRINT.**

| | Simulations | | | | | | | | | | Original |
|---|---|---|---|---|---|---|---|---|---|---|---|
| | #1 | #2 | #3 | #4 | #5 | #6 | #7 | #8 | #9 | #10 | |
| **Total number** | 6844 | 7854 | 7990 | 5896 | 6525 | 7990 | 9075 | 7581 | 9361 | 7990 | 9361 |
| **Intensive arm** | 3485 (50.9) | 3893 (49.6) | 3992 (50.0) | 2926 (49.6) | 3222 (49.4) | 3998 (50.0) | 4526 (49.9) | 3770 (49.7) | 4678 (50.0) | 3978 (49.8) | 4678 (50.0) |
| **Age** | 68.2 (9.4) | 67.8 (9.4) | 68.0 (9.4) | 67.8 (9.4) | 67.6 (9.4) | 67.9 (9.4) | 67.9 (9.4) | 67.6 (9.4) | 67.9 (9.4) | 68.0 (9.4) | 67.9 (9.4) |
| **Female sex** | 2297 (33.6) | 2861 (36.4) | 2826 (35.4) | 2224 (37.7) | 2098 (32.2) | 2756 (34.5) | 3209 (35.4) | 2790 (36.8) | 3332 (35.6) | 2868 (35.9) | 3332 (35.6) |
| ***Non-Hispanic Black*** | 1967 (28.7) | 2383 (30.3) | 2342 (29.3) | 1924 (32.6) | 2000 (30.7) | 2390 (29.9) | 2758 (30.4) | 2353 (31.0) | 2802 (29.9) | 2400 (30.0) | 2802 (29.9) |
| ***Hispanic*** | 721 (10.5) | 851 (10.8) | 850 (10.6) | 630 (10.7) | 634 (9.7) | 853 (10.7) | 946 (10.4) | 828 (10.9) | 984 (10.5) | 841 (10.5) | 984 (10.5) |
| ***Non-Hispanic White*** | 4016 (58.7) | 4467 (56.9) | 4645 (58.1) | 3223 (54.7) | 3752 (57.5) | 4593 (57.5) | 5202 (57.3) | 4242 (56.0) | 5399 (57.7) | 4591 (57.5) | 5399 (57.7) |
| **BMI (kg/m$^2$)** | 29.8 (5.7) | 29.8 (5.8) | 29.8 (5.7) | 29.9 (5.8) | 29.9 (5.7) | 29.8 (5.7) | 29.8 (5.7) | 29.9 (5.8) | 29.8 (5.8) | 29.8 (5.7) | 29.8 (5.8) |
| **Blood pressure medications** | 2.0 [1.0,3.0] | 2.0 [1.0,3.0] | 2.0 [1.0,3.0] | 2.0 [1.0,3.0] | 2.0 [1.0,3.0] | 2.0 [1.0,3.0] | 2.0 [1.0,3.0] | 2.0 [1.0,3.0] | 2.0 [1.0,3.0] | 2.0 [1.0,3.0] | 2.0 [1.0,3.0] |
| ***Former*** | 3138 (45.9) | 3321 (42.3) | 3510 (43.9) | 2312 (39.2) | 2646 (40.6) | 3521 (44.1) | 3896 (42.9) | 3006 (39.7) | 3983 (42.5) | 3385 (42.4) | 3983 (42.5) |
| ***Active*** | 797 (11.6) | 1060 (13.5) | 992 (12.4) | 720 (12.2) | 841 (12.9) | 1079 (13.5) | 1207 (13.3) | 1002 (13.2) | 1242 (13.3) | 1014 (12.7) | 1242 (13.3) |
| **Aspirin use** | 3917 (57.2) | 3925 (50.0) | 4012 (50.2) | 2971 (50.4) | 3278 (50.2) | 4038 (50.5) | 4573 (50.4) | 3472 (45.8) | 4772 (51.0) | 4170 (52.2) | 4772 (51.0) |
| **Statin use** | 3372 (49.3) | 3402 (43.3) | 3489 (43.7) | 2491 (42.2) | 2807 (43.0) | 3526 (44.1) | 3962 (43.7) | 2934 (38.7) | 4080 (43.6) | 3439 (43.0) | 4080 (43.6) |
| **Cardiovascular disease** | 1693 (24.7) | 1621 (20.6) | 1596 (20.0) | 1070 (18.1) | 966 (14.8) | 1605 (20.1) | 1846 (20.3) | 1244 (16.4) | 1877 (20.1) | 1618 (20.3) | 1877 (20.1) |
| **Chronic kidney disease** | 1758 (25.7) | 2054 (26.2) | 2259 (28.3) | 1541 (26.1) | 1582 (24.2) | 2243 (28.1) | 2561 (28.2) | 2051 (27.1) | 2646 (28.3) | 2258 (28.3) | 2646 (28.3) |
| **Myocardial infarction** | 606 (8.9) | 578 (7.4) | 568 (7.1) | 370 (6.3) | 339 (5.2) | 571 (7.1) | 659 (7.3) | 435 (5.7) | 666 (7.1) | 577 (7.2) | 666 (7.1) |
| **Coronary revascularization** | 806 (11.8) | 753 (9.6) | 741 (9.3) | 457 (7.8) | 431 (6.6) | 750 (9.4) | 868 (9.6) | 560 (7.4) | 878 (9.4) | 754 (9.4) | 878 (9.4) |
| **Peripheral vascular disease** | 414 (6.0) | 415 (5.3) | 416 (5.2) | 296 (5.0) | 304 (4.7) | 419 (5.2) | 482 (5.3) | 357 (4.7) | 503 (5.4) | 421 (5.3) | 503 (5.4) |
| **Carotid artery disease** | 271 (4.0) | 253 (3.2) | 252 (3.2) | 160 (2.7) | 154 (2.4) | 243 (3.0) | 291 (3.2) | 192 (2.5) | 298 (3.2) | 251 (3.1) | 298 (3.2) |
| **Framingham risk score** | 17.3 (2.5) | 17.4 (2.5) | 17.4 (2.5) | 17.3 (2.5) | 17.4 (2.5) | 17.4 (2.5) | 17.4 (2.5) | 17.4 (2.5) | 17.4 (2.5) | 17.4 (2.5) | 17.4 (2.5) |
| **Atrial fibrillation** | 566 (8.3) | 611 (7.8) | 658 (8.2) | 503 (8.5) | 490 (7.5) | 618 (7.7) | 722 (8.0) | 594 (7.8) | 754 (8.1) | 657 (8.2) | 754 (8.1) |
| **Congestive heart failure** | 252 (3.7) | 274 (3.5) | 277 (3.5) | 212 (3.6) | 209 (3.2) | 269 (3.4) | 321 (3.5) | 250 (3.3) | 326 (3.5) | 287 (3.6) | 326 (3.5) |
| **Family history** | 4247 (62.1) | 4903 (62.4) | 5090 (63.7) | 3723 (63.1) | 3994 (61.2) | 4924 (61.6) | 5808 (64.0) | 4726 (62.3) | 6028 (64.4) | 5035 (63.0) | 6028 (64.4) |
| **SBP (mmHg)** | 139.6 (15.7) | 139.6 (15.5) | 139.5 (15.4) | 139.7 (15.6) | 139.8 (15.5) | 139.5 (15.4) | 139.7 (15.5) | 140.0 (15.7) | 139.7 (15.6) | 139.7 (15.6) | 139.7 (15.6) |
| **DBP (mmHg)** | 77.8 (11.9) | 78.2 (11.9) | 78.1 (11.8) | 78.3 (11.9) | 78.6 (11.8) | 78.1 (11.8) | 78.1 (12.0) | 78.6 (11.9) | 78.1 (11.9) | 78.1 (11.9) | 78.1 (11.9) |
| **Resting heart rate (bpm)** | 65.9 (11.5) | 66.4 (11.6) | 66.3 (11.6) | 66.3 (11.5) | 66.4 (11.6) | 66.3 (11.6) | 66.3 (11.6) | 66.5 (11.6) | 66.3 (11.6) | 66.2 (11.5) | 66.3 (11.6) |
| **Total cholesterol (mg/dL)** | 188.2 (41.1) | 190.3 (41.2) | 189.9 (41.2) | 191.2 (40.6) | 190.5 (40.0) | 189.9 (41.2) | 190.1 (41.1) | 192.2 (40.9) | 190.1 (41.1) | 190.0 (41.1) | 190.1 (41.1) |
| **HDL (mg/dL)** | 52.6 (14.0) | 53.0 (14.5) | 53.0 (14.6) | 53.2 (14.6) | 52.8 (14.1) | 52.7 (14.5) | 52.9 (14.5) | 53.1 (14.7) | 52.9 (14.4) | 53.0 (14.5) | 52.9 (14.4) |
| **LDL (mg/dL)** | 111.1 (34.9) | 112.6 (35.1) | 112.3 (35.1) | 113.5 (34.8) | 113.2 (34.1) | 112.4 (35.1) | 112.6 (35.1) | 114.4 (34.9) | 112.6 (35.1) | 112.4 (35.0) | 112.6 (35.1) |
| **eGFR (mL/min/1.73m$^2$)** | 72.5 (20.3) | 72.6 (20.3) | 71.8 (20.5) | 72.5 (20.4) | 73.3 (20.2) | 71.9 (20.6) | 71.8 (20.6) | 72.3 (20.5) | 71.8 (20.6) | 71.8 (20.4) | 71.8 (20.6) |

BMI: body mass index; DBP: diastolic blood pressure; eGFR: estimated glomerular filtration rate; HDL: High-density lipoprotein; LDL: Low-density lipoprotein; SBP: systolic blood pressure. Continuous variables are summarized as mean (standard deviation) or median [25th-75th percentile], categorical variables as counts (valid percentages). All variables presented had <10% missing data at baseline.

**Supplementary Table 6. Key parameters, rationale, and potential implications.**

| Feature | Default | Rationale | Implications for deployment |
|---|---|---|---|
| **Split ratio to create trial phenomaps** | 50:50 | A 50:50 split maximizes the ability of the training set to detect possible patterns of heterogeneity of the treatment effect, while allowing the testing set to test the performance of the trained regressor. Furthermore, the equal sizes of the two samples (training and testing set) balances the discovery and validation of treatment effect heterogeneity and minimize variability in the distance metrics that may arise because of imbalanced dataset splits. | The choice of this parameter can modified depending on the anticipated size of the study or rate of expected events. However, given the reliance of the model on both discovery and validation of a heterogeneous effect signal, it is recommended that the two sets remain balanced in size. |
| **Split ratio to train the XGBoost regressor in the training set** | 80:20 | Given that the regressor is trained against continuous values, the size of the training set is prioritized to create a reliable model, which will be more likely to detect meaningful heterogeneity in the testing set. | This can be adapted further as an iterative process, finding the optimal split that results in the most consistent effects in the testing set. |
| ***p* value for interaction** | 0.2 | Given that these analyses are performed during interim timepoints across randomly selected observation subgroups, a p value for interaction that is higher than the commonly used 0.05 or 0.1 cutoff may be needed. Higher values will enable greater flexibility to discover early signals of heterogeneity, with the caveat that it may make the algorithm more sensitive to false discoveries and noise. A lower value would make the algorithm more conservative. | Given that most studies are not powered to detect heterogeneous treatment effects, this threshold reflects a balance between screening for an early heterogeneous effect, while reducing the risk of false discovery.<br>**Regulatory implications:** This level may need to be reviewed by key stakeholders during the design of the protocol and adjusted to a lower value to maximize the reliability of any discovered signals. |
| Winsorization | 0.95 | The presence of outliers may have disproportionate impact on the definition of the phenomaps; to avoid this, winsorization of continuous variables is recommended by winsorizing outliers above the 97.5th percentile and below the 2.5th percentile to the respective values of these percentiles. | This is often required in real-world data to avoid the effect of outliers and prevent false discoveries driven by unusual data distributions. |
| **Dissimilarity matrix distance** | Gower's | Gower's distance enables the processing of mixed datasets, which are common in clinical trial research. Alternative distances may be used at the discretion of the investigators. | This can be optimized to the specific nature of each dataset. In the case of matrices of continuous variables, alternative distance metrics (i.e., Euclidean) may be preferred. |
| **Sigmoid transformation of probabilities to guide enrollment** | Sigmoid | Any sigmoid, linear, or other transformation may be applied to ensure higher probabilities of enrollment among those predicted as higher responders. A sigmoid transformation was chosen here to ensure a monotonic association between higher predicted response and higher probability of enrollment with a plateau zone for the highest and lowest responders. | Regulatory implications: This directly affects the enrichment process and needs to be agreed upon during the protocol stage. |
| **Number of interim analyses and their pre-defined timepoints** | 3 | The number of interim analyses and their pre-defined timepoints along the course of the trial should be reviewed depending on the size, nature and expected length of a clinical trial. | More analyses offer the opportunity to define or revise estimates of heterogeneous effects, but should be weighed against the statistical (i.e., alpha-spending) and practical cost. In pragmatic (e.g., electronic health record-embedded) trials this could be automated and run in real-time, with the results made available to the statistical review team at each pre-defined timepoint. |

XGBoost: extreme gradient boosting.

#### Supplementary Figures

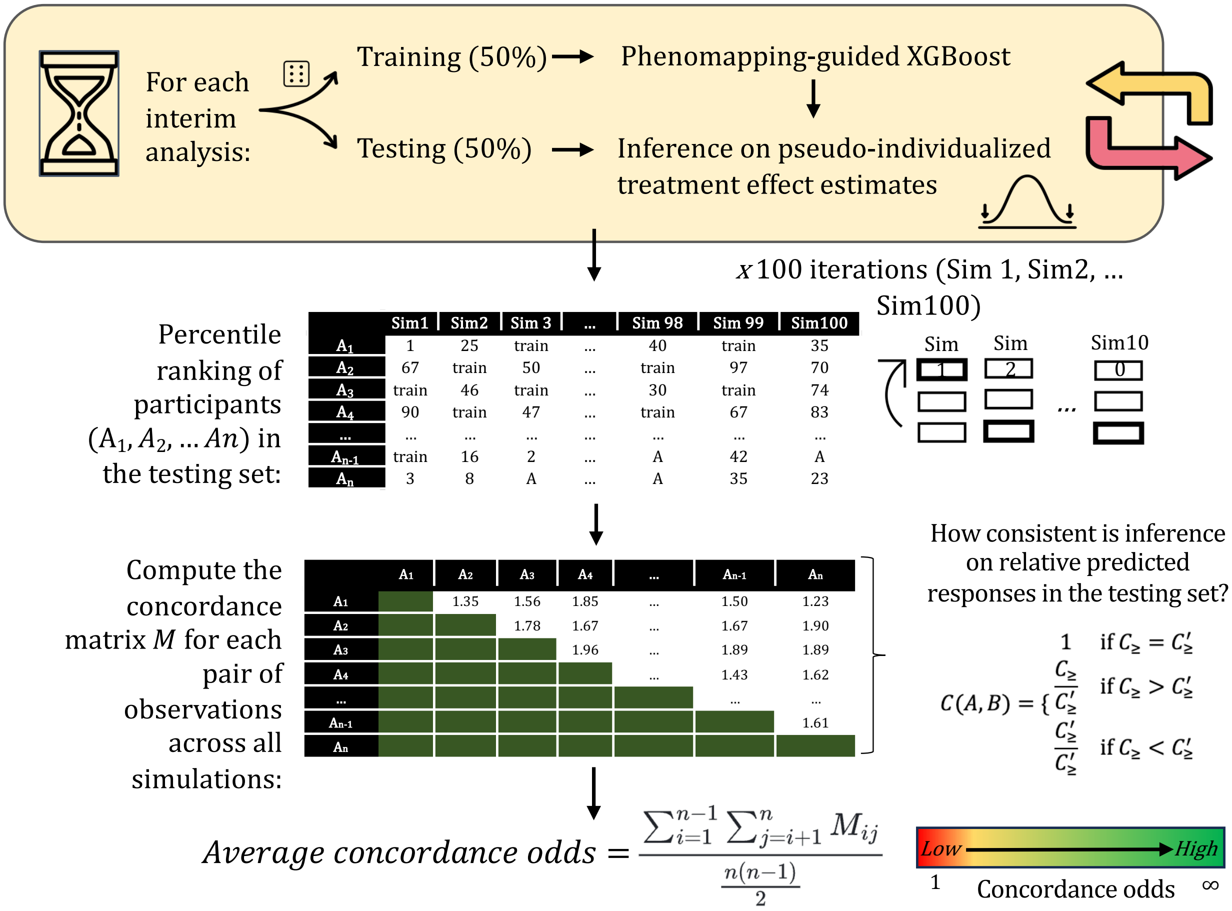


**Supplementary Figure 1 | Stability of relative predicted treatment effect.** To better understand the stability of the predicted response ranks, we randomly split each study trial population at the time of each interim analysis timepoint across 100 iterations. Within each iteration, we ranked individuals in the testing set based on their predicted treatment response as defined based on the extreme gradient boosting regressor trained in the training set. We then calculated a concordance metric/ratio of the relative ranks for a random combination of patients A and B, when both patients were present in the testing set. Averaging this metric for all unique patient combinations yielded an average concordance ratio, with higher values (>1) reflecting greater stability in the respective ranks and treatment response profile. XGBoost: extreme gradient boosting.

**Example of model evolution during one simulation in IRIS**

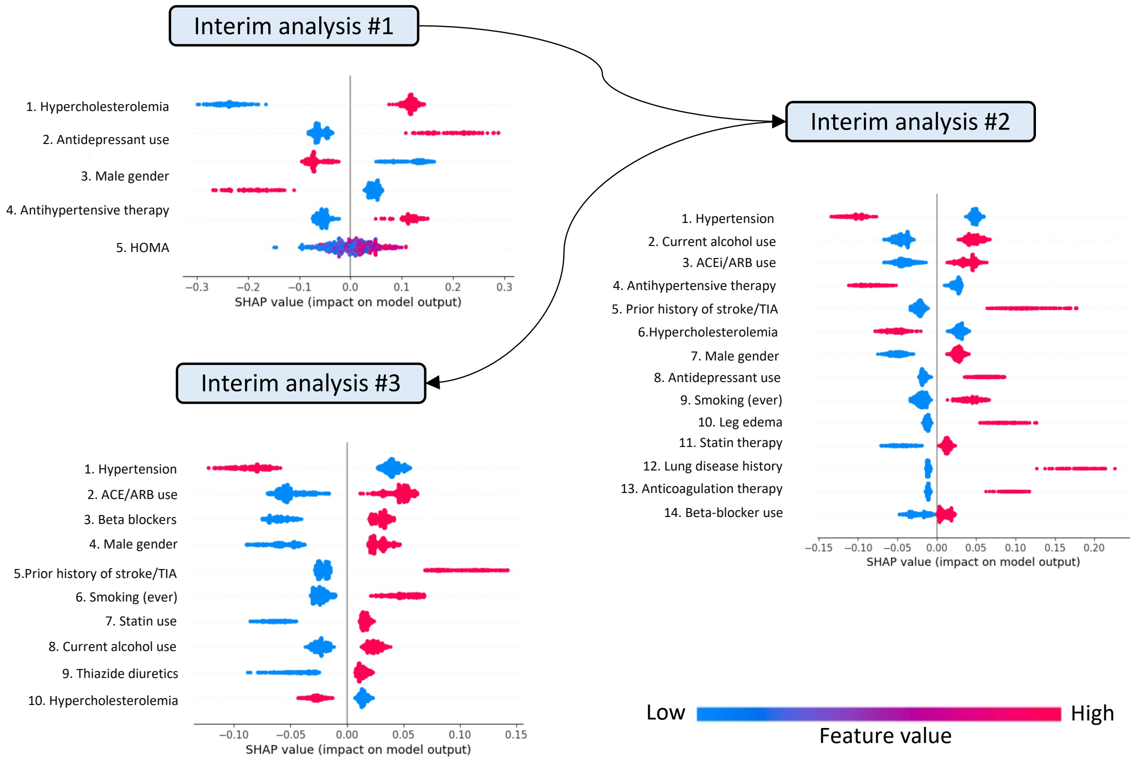


**Supplementary Figure 2 | Example of model evolution for prediction of the personalized cardiovascular benefit of pioglitazone versus placebo across different enrollment periods.** The panels show the SHAP (Shapley additive explanations) plot summary for the final epoch model using the features identified as important by the Boruta method. The features are ranked from most to least important based on their SHAP values. Negative SHAP values favor the intensive systolic blood pressure reduction. Red color denotes higher numerical values for a given feature, or in the case of categorical features, the presence of that feature. Blue color denotes lower numerical values or the absence of a categorical feature. ACEi: Angiotensin-converting-enzyme inhibitors; ARB: angiotensin receptor blockers; IRIS: Insulin Resistance Intervention after Stroke; TIA: transient ischemic attack.

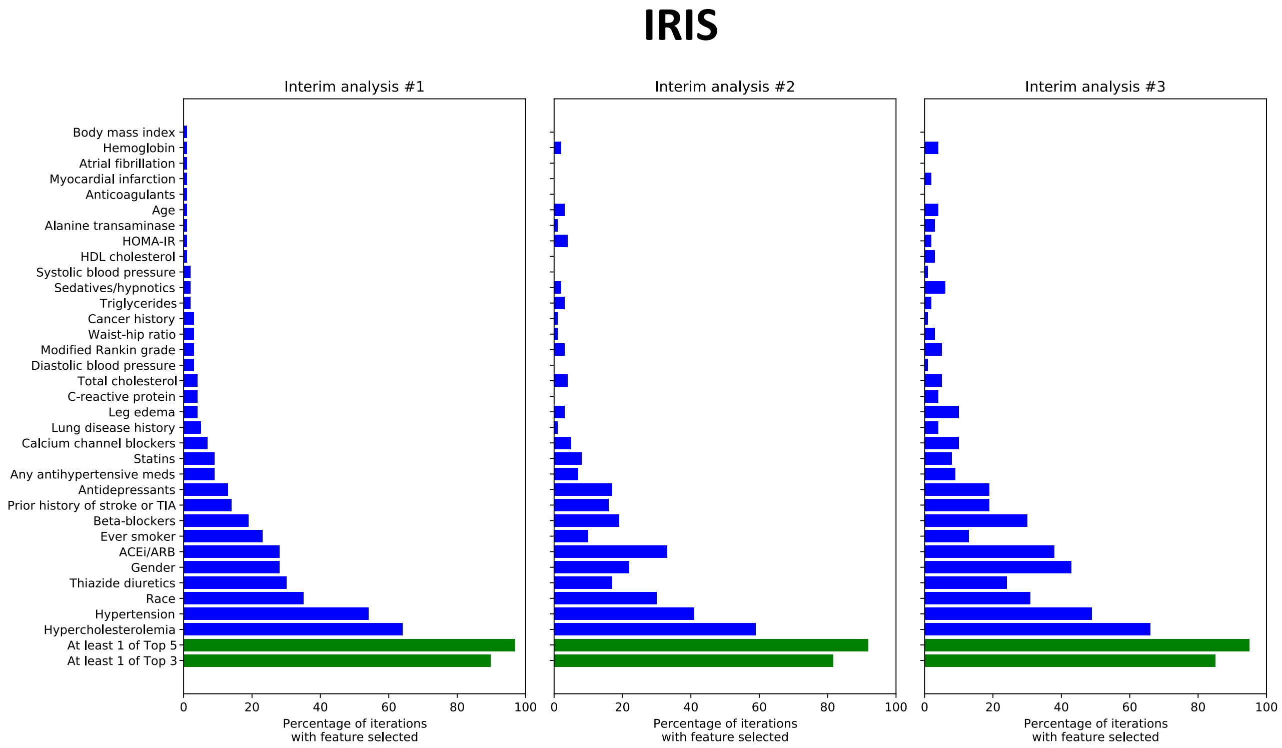


**Supplementary Figure 3 | Stability of predictors in IRIS.** Selection frequency of individual features based on the Boruta SHAP algorithm across the three interim analysis timepoints in the IRIS trial. Blue bars refer to individual features, whereas green bars denote the presence of at least one of the top 3 or top 5 predictors identified during the first interim analysis (left panel). ACEi: Angiotensin-converting-enzyme inhibitors; ARB: angiotensin receptor blockers; HDL: High-density lipoprotein; HOMA-IR: Homeostatic Model Assessment for Insulin Resistance; IRIS: Insulin Resistance Intervention after Stroke; TIA: transient ischemic attack.

**Example of model evolution during one simulation in SPRINT**

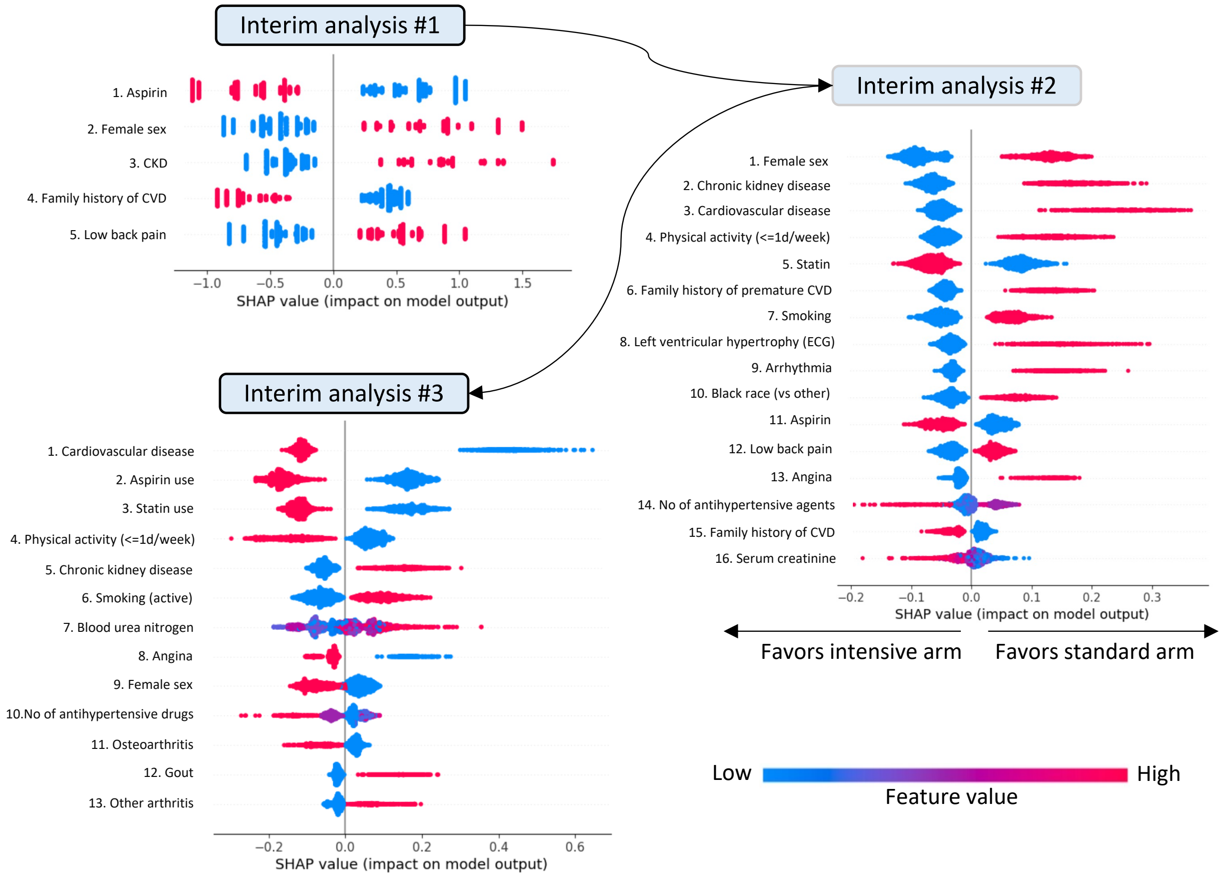


**Supplementary Figure 4 | Example of model evolution for prediction of the personalized cardiovascular benefit of intensive versus standard systolic blood pressure reduction across different enrollment periods.** The panels show the SHAP (Shapley additive explanations) plot summary for the final epoch model using the features identified as important by the Boruta method. The features are ranked from most to least important based on their SHAP values. Negative SHAP values favor the intensive systolic blood pressure reduction. Red color denotes higher numerical values for a given feature, or in the case of categorical features, the presence of that feature. Blue color denotes lower numerical values or the absence of a categorical feature. CVD: cardiovascular disease; CKD: chronic kidney disease; SPRINT: Systolic Blood Pressure Intervention Trial.

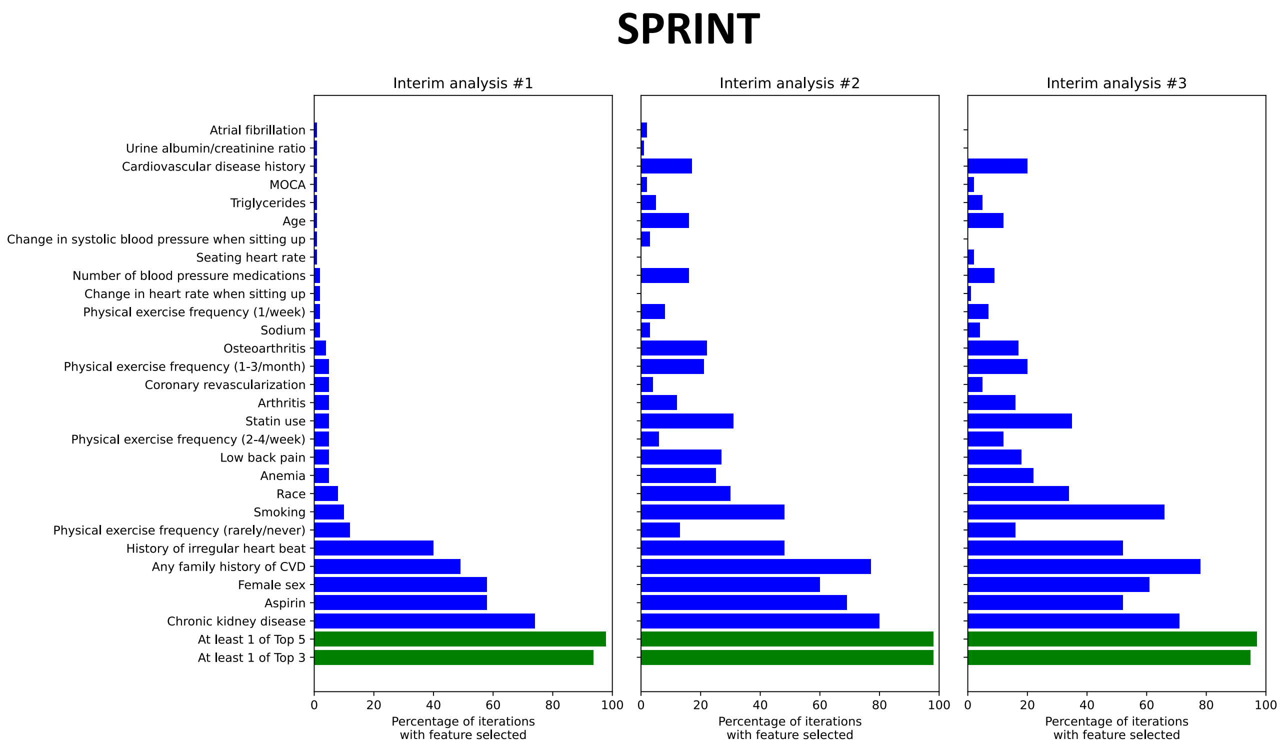


**Supplementary Figure 5 | Stability of predictors in SPRINT.** Selection frequency of individual features based on the Boruta SHAP algorithm across the three interim analysis timepoints in SPRINT. Blue bars refer to individual features, whereas green bars denote the presence of at least one of the top 3 or top 5 predictors identified during the first interim analysis (left panel). CVD: cardiovascular disease; MoCA: Montreal Cognitive Assessment ; SPRINT: Systolic Blood Pressure Intervention Trial.

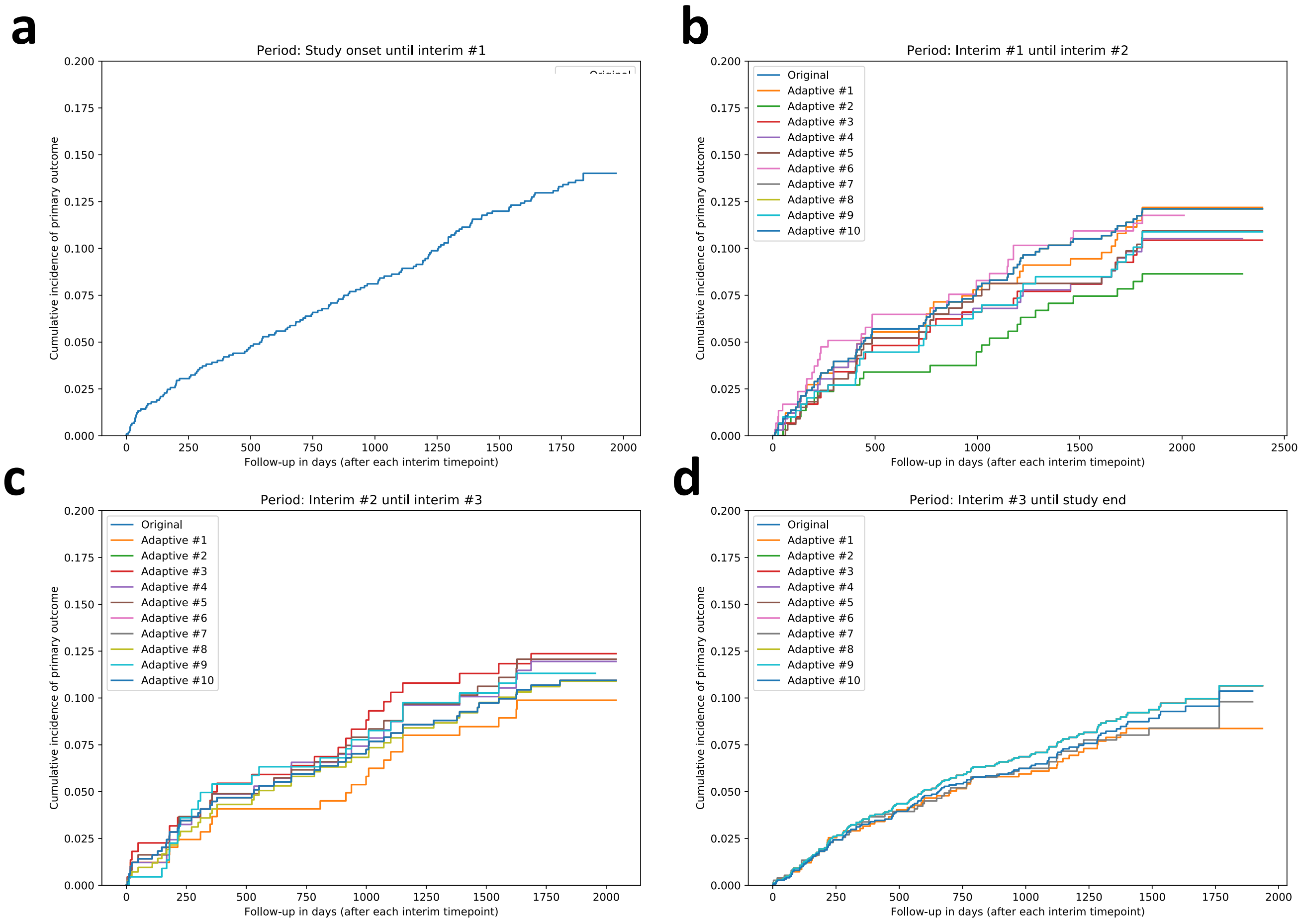


**Supplementary Figure 6 | Incidence of the primary outcome across different enrollment periods in IRIS.** Incidence of the primary outcome among patients recruited: **(A)** prior to the first interim analysis, **(B)** between the first and second interim timepoints, **(C)** second and third timepoints, and **(D)** after the third interim analysis timepoint. Each analysis includes distinct curves for each one of the adaptive simulations. IRIS: Insulin Resistance Intervention after Stroke.

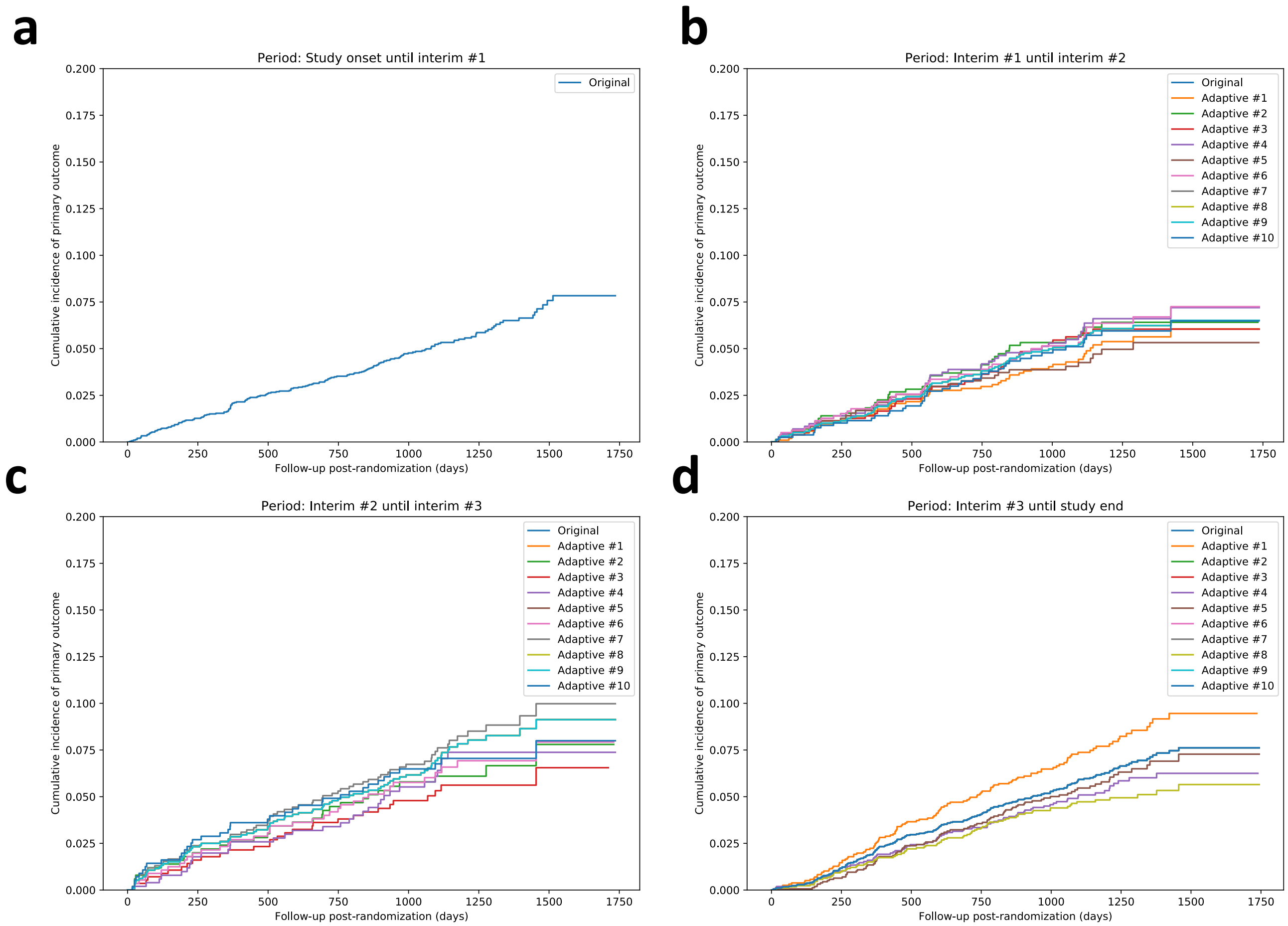


**Supplementary Figure 7 | Incidence of the primary outcome across different enrollment periods in SPRINT.** Incidence of the primary outcome among patients recruited: (A) prior to the first interim analysis, (B) between the first and second interim timepoints, (C) second and third timepoints, and (D) after the third interim analysis timepoint. Each analysis includes distinct curves for each one of the adaptive simulation**s**. SPRINT: Systolic Blood Pressure Intervention Trial.
